# Comparative Mortality Risk of Aripiprazole, Olanzapine, Quetiapine and Risperidone in Alzheimer’s Disease: A Real□World Cohort Study with Treatment Effect Heterogeneity Analysis

**DOI:** 10.1101/2025.11.13.25340096

**Authors:** Chen Jiang, Josh Krivinko, Zeshui Yu, Robert A. Sweet, Lang Zeng, Hui Wang, Ying Ding, Zheng Zeng, Julia Kofler, Lirong Wang

## Abstract

**Background:** Second-generation antipsychotics (SGAs) are frequently used off-label to manage behavioral symptoms in Alzheimer’s disease (AD), despite ongoing concerns about their safety. Comparative evidence on mortality risk across specific SGAs remains limited.

**Objective:** To compare all-cause mortality among AD patients treated with commonly prescribed SGAs and to explore treatment effect heterogeneity using causal machine learning.

**Methods:** We conducted a retrospective cohort study using de-identified electronic health records from the Truveta platform (2018–2024). Patients with incident AD initiating treatment with aripiprazole, risperidone, quetiapine, or olanzapine were identified using an active-comparator, new-user design. Drug exposure was modeled as a time-varying covariate in Cox proportional hazards models, with propensity score matching applied to control for confounding. Causal tree and targeted maximum likelihood estimation (TMLE) were used to identify subgroups with heterogeneous treatment effects.

**Results:** Among 17,004 AD patients, aripiprazole was associated with significantly lower mortality than olanzapine (HR = 0.667, 95% CI: 0.472–0.941) and quetiapine (HR = 0.677, 95% CI: 0.462–0.990). Quetiapine was also associated with lower mortality than olanzapine (HR = 0.833, 95% CI: 0.702–0.990) and risperidone (HR = 0.830, 95% CI: 0.705–0.978). Causal tree analysis revealed treatment effect heterogeneity by clinical characteristics, particularly among patients using type 2 diabetes (T2DM) medications. In subgroup analyses, aripiprazole remained protective in T2DM users (HR = 0.604 vs. quetiapine and risperidone, p = 0.002).

**Conclusions:** Mortality risks vary substantially across SGAs in AD patients. Aripiprazole and quetiapine were associated with lower mortality compared to olanzapine and risperidone. Treatment effect heterogeneity suggests the need for individualized prescribing based on patient characteristics such as comorbid T2DM.

## Introduction

Alzheimer’s disease (AD) is the most common form of dementia and is frequently accompanied by behavioral and psychological symptoms, including psychosis[1]. These symptoms often lead clinicians to prescribe second□generation antipsychotics (SGAs) off□label in this vulnerable population[2]. Due to findings of increased risks of cerebrovascular events and cardiovascular-related deaths associated with SGA use in older adults with dementia[3], the U.S. Food and Drug Administration (FDA) to issue a “black box warning” for these medications in 2005[4]. Since then, concerns about the safety of SGAs in treating dementia-related psychosis have persisted.

Although concerns about antipsychotic safety in dementia have been widely studied, most existing research has focused on comparisons between antipsychotic use and placebo or non-use[5, 6]. While these studies have been informative, such designs do not allow for the evaluation of relative risks across different SGAs. Direct head-to-head comparisons of SGAs in patients with AD, particularly with respect to all-cause mortality remain limited. In clinical practice, after carefully weighing the risks and benefits of initiating antipsychotic treatment, and when treatment is deemed necessary, clinicians must choose among several antipsychotic agents. Therefore, comparative safety profiles of individual SGAs are of critical importance to guide prescribing decisions in real-world settings. Moreover, despite the widespread use of SGAs, there is limited research using causal machine learning methods to explore their treatment effect heterogeneity. Given the complex comorbidities and medication profiles often seen in patients with AD[7], identifying subgroups that may gain greater benefit or face greater harm from specific SGAs is essential to inform personalized treatment strategies.

To address these knowledge gaps, we conducted a real-world cohort study of patients with AD undergoing treatment with commonly prescribed SGAs (aripiprazole, risperidone, quetiapine and olanzapine). We employed an active-comparator, new-user design and time-varying Cox regression to estimate all-cause mortality risk across 4 specific SGAs. In addition, we applied treatment effect heterogeneity analysis using causal tree and targeted maximum likelihood estimation to identify clinically meaningful subgroups that may exhibit heterogeneous responses to different SGAs. This study not only extends existing safety data on antipsychotic use, but also contributes new evidence to support individualized treatment strategies in AD population.

## Method

### Design and Data Source

We conducted a retrospective cohort study using de-identified electronic health record (EHR) data from the Truveta platform, covering the period from 2018 to 2024. The Truveta platform aggregates longitudinal, de-identified EHR data from participating health systems across the United States, representing more than 120 million patients. The database integrates structured clinical information, including demographics, diagnoses, procedures, medication dispensing, vital signs, and laboratory results. This retrospective cohort study followed the Strengthening the Reporting of Observational Studies in Epidemiology (STROBE) reporting guideline[8].

### Cohort selection

The study cohort included adults with incident Alzheimer’s disease (AD) prior to SGAs treatment initiation, defined as patients who received a new AD diagnosis after at least one year of documented healthcare activity in the Truveta data network and had no prior AD diagnosis. AD was identified using at least one diagnostic code from ICD-10-CM (G30 and its descendants), ICD-9-CM (331.0), or SNOMED CT (26929004 and its descendants). To ensure adequate longitudinal data, eligible patients were required to have at least one valid healthcare encounter recorded ≥ 365 days before the first AD diagnosis and at least one additional encounter within 1–365 days after diagnosis.

Patients also had to have at least one follow-up encounter after treatment initiation to allow estimation of survival time.

Patients were excluded if they received both drugs under comparison during the study period (for example, individuals who used both aripiprazole and olanzapine were excluded when we were comparing aripiprazole and olanzapine). Patients with death date record before the last dispensing date were also excluded.

### Exposure

Exposure was defined based on the dispense record of four second-generation antipsychotics (aripiprazole, olanzapine, quetiapine, and risperidone) that are the most frequently used SGAs in the study cohort. We defined the index date as the date of the first prescription dispensing of a target SGA after the first AD diagnosis. To approximate a new-user design, patients with any record of SGA use in the 180 days prior to the index date were excluded. Six head-to-head treatment comparisons were constructed among these treatments. Additionally, exposure definitions were mutually exclusive across comparison pairs.

Drug exposure was modeled as a time-varying variable. For each individual, dispensing dates and days of supply were used to define the start and end of each exposure period. Consecutive exposure separated by less than 30 days were regarded as one continuous exposure window, and gaps more than 30 days were considered the start of a new episode[9, 10]. Each patient’s follow-up timeline was divided into discrete intervals corresponding to periods of exposure and non-exposure. Exposure status was formatted dynamically within these intervals.

### Covariates and Confounding Control

Covariate selection was informed by clinical relevance, prior evidence of associations with mortality, and the need to capture heterogeneity in treatment effects. The propensity score model and subsequent heterogeneity analyses incorporated demographic characteristics (age, sex, race, and marital status); comorbidities including heart failure, myocardial infarction, stroke, chronic kidney disease, chronic obstructive pulmonary disease, type 2 diabetes mellitus, cancer, hypertension, substance use disorders, and depression; and concurrent use of medications commonly prescribed medications in AD patients, such as FDA-approved medications for AD, antihypertensive, antidiabetic, lipid-lowering, antidepressant, benzodiazepine[11, 12], anticholinergic[13], and analgesic agents[11, 14].

To preserve data granularity for causal tree analyses, individual comorbidities were modeled as separate binary covariates rather than summarized using composite index such as the Charlson or Elixhauser Comorbidity Index. This was critical because composite indices can obscure the specific effects of individual conditions, which is essential for detecting heterogeneity. Our approach allowed the exploration of potential treatment effect heterogeneity across specific clinical characteristics while maintaining adequate confounding control in propensity score estimation.

### Outcome

The primary outcome was all-cause mortality. Deaths were identified from death records documented in TRUVETA’s EHR system. Survival time was calculated as the number of days from the index prescription dispense date to death or censoring. Deaths that occurred during an active dispensing period or within 30 days after the end of the last exposure episode were classified as events. Patients who died beyond this 30-day window, were lost to follow-up (no encounter after the exposure window plus 30 days), or remained alive at 2 years post-index were treated as censored.

### Statistical Analysis

Propensity score matching (PSM) was conducted using the *MatchIt[15]* package in R (version 4.2.0), applying nearest-neighbor matching with a 1:1 ratio. All previously described covariates were included in the propensity score model. Covariate balance before and after matching was assessed using standardized mean differences (SMDs). SMD less than 0.1 indicated a balanced matching result. The matched datasets were then used for subsequent survival analyses and treatment heterogeneity analyses.

We used time-varying Cox proportional hazards models for survival analysis, with treatment exposure incorporated as a time-dependent covariate. The outcome of interest was the time from the index date to death or censoring. The event definition was consistent with the primary outcome specification, considering only deaths occurring within 30 days after the end of the last exposure period. Follow-up period was truncated at two years to maintain sufficient sample size and stability of the hazard ratio estimates. For each drug comparison, hazard ratios (HRs) and 95% confidence intervals (CIs) were reported, and results were summarized graphically using forest plots.

To investigate potential heterogeneity in treatment effects, *CausalTree* package in R was applied to the matched dataset to identify patient subgroups with differential mortality effects[16]. For each terminal node, the average treatment effect (ATE) and corresponding 95% CI were estimated using targeted maximum likelihood estimation (TMLE) with *tmle[17]* and *SuperLearner[18]* package in R. Subgroups whose 95% CIs did not cross zero were interpreted as showing statistically significant treatment effect heterogeneity. The causal tree structures and node-specific estimates were visualized to illustrate the patterns of heterogeneity across patient subgroups.

## Result

### Characteristics of the Study Cohort

A total of 17,004 patients with Alzheimer’s disease were included across six head-to-head drug comparison cohorts. **Table 1** shows the baseline characteristics of patients treated with aripiprazole and olanzapine before and after propensity score matching (PSM). Prior to matching, patients receiving aripiprazole were younger (mean age: 80.11 vs. 83.02 years) and had higher rates of depression compared with olanzapine users (61.6% vs. 42.2%). After 1:1 matching, 790 patients remained in each treatment group, and baseline characteristics were well-balanced across all measured variables (absolute standardized mean differences < 0.05). The matched cohort had a mean age of approximately 80 years old, 70% of the cohort were female, and the majority were White (∼90%). Patients had frequent comorbidities, with common conditions including hypertension (∼72%), depression (∼61%), type 2 diabetes mellitus (∼30%), and chronic kidney disease (∼28%). More than half of the patients were receiving antidepressants, and approximately half were using anti□dementia medications at baseline. Baseline characteristics for the remaining five drug comparison pairs are provided in **Supplementary Tables S1–S5**.

**Table 1.**
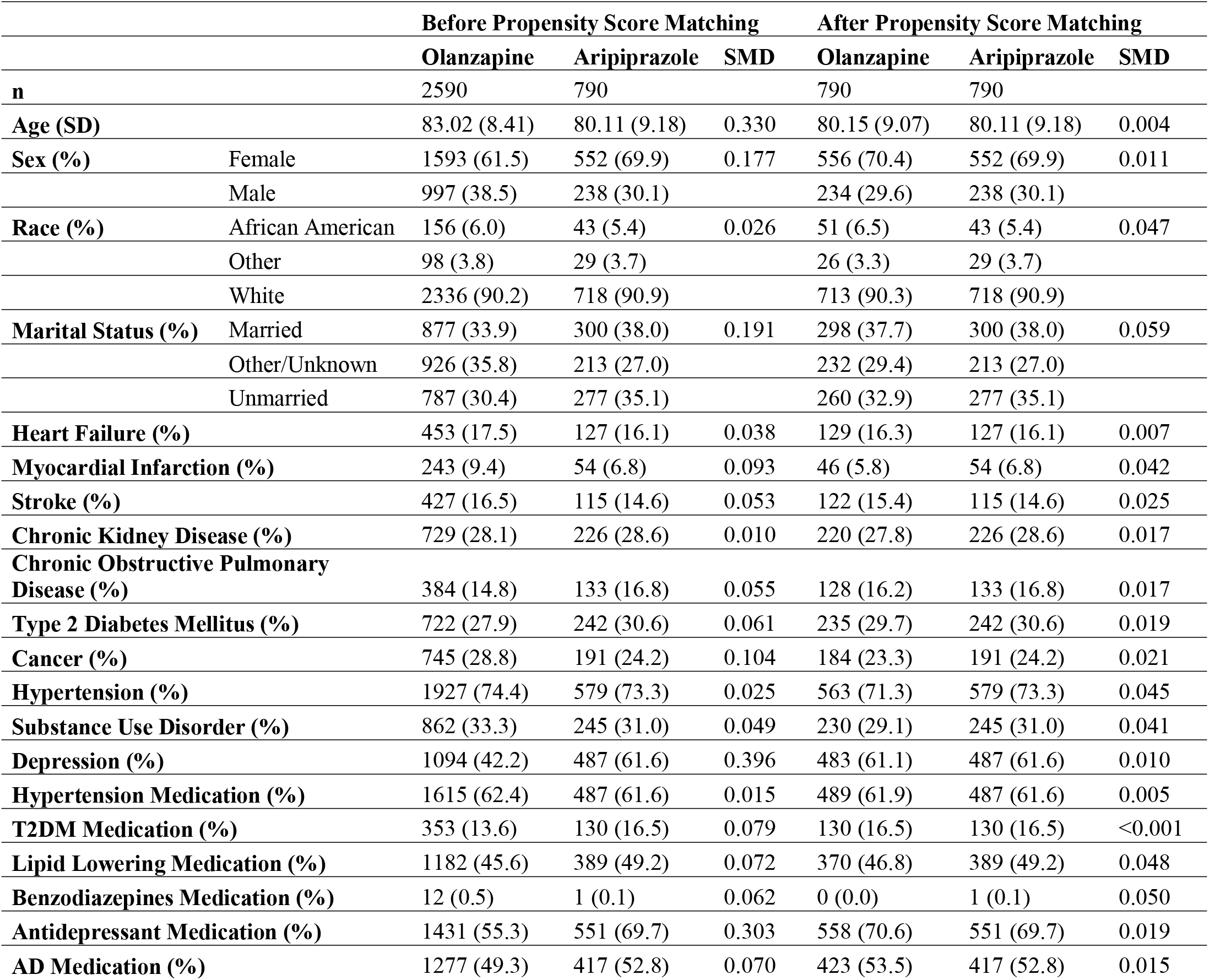
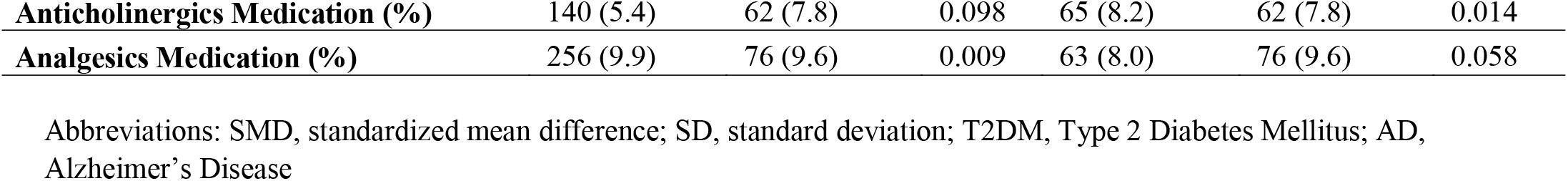
Baseline Characteristics of Patients Treated with Aripiprazole and Olanzapine Before and After Propensity Score Matching.

|  |  | Before Propensity Score Matching |  |  | After Propensity Score Matching |  |  |
| --- | --- | --- | --- | --- | --- | --- | --- |
|  |  | Olanzapine | Aripiprazole | SMD | Olanzapine | Aripiprazole | SMD |
| **n** |  | 2590 | 790 |  | 790 | 790 |  |
| **Age (SD)** |  | 83.02 (8.41) | 80.11 (9.18) | 0.330 | 80.15 (9.07) | 80.11 (9.18) | 0.004 |
| **Sex (%)** | Female | 1593 (61.5) | 552 (69.9) | 0.177 | 556 (70.4) | 552 (69.9) | 0.011 |
|  | Male | 997 (38.5) | 238 (30.1) |  | 234 (29.6) | 238 (30.1) |  |
| **Race (%)** | African American | 156 (6.0) | 43 (5.4) | 0.026 | 51 (6.5) | 43 (5.4) | 0.047 |
|  | Other | 98 (3.8) | 29 (3.7) |  | 26 (3.3) | 29 (3.7) |  |
|  | White | 2336 (90.2) | 718 (90.9) |  | 713 (90.3) | 718 (90.9) |  |
| **Marital Status (%)** | Married | 877 (33.9) | 300 (38.0) | 0.191 | 298 (37.7) | 300 (38.0) | 0.059 |
|  | Other/Unknown | 926 (35.8) | 213 (27.0) |  | 232 (29.4) | 213 (27.0) |  |
|  | Unmarried | 787 (30.4) | 277 (35.1) |  | 260 (32.9) | 277 (35.1) |  |
| **Heart Failure (%)** |  | 453 (17.5) | 127 (16.1) | 0.038 | 129 (16.3) | 127 (16.1) | 0.007 |
| **Myocardial Infarction (%)** |  | 243 (9.4) | 54 (6.8) | 0.093 | 46 (5.8) | 54 (6.8) | 0.042 |
| **Stroke (%)** |  | 427 (16.5) | 115 (14.6) | 0.053 | 122 (15.4) | 115 (14.6) | 0.025 |
| **Chronic Kidney Disease (%)** |  | 729 (28.1) | 226 (28.6) | 0.010 | 220 (27.8) | 226 (28.6) | 0.017 |
| **Chronic Obstructive Pulmonary Disease (%)** |  | 384 (14.8) | 133 (16.8) | 0.055 | 128 (16.2) | 133 (16.8) | 0.017 |
| **Type 2 Diabetes Mellitus (%)** |  | 722 (27.9) | 242 (30.6) | 0.061 | 235 (29.7) | 242 (30.6) | 0.019 |
| **Cancer (%)** |  | 745 (28.8) | 191 (24.2) | 0.104 | 184 (23.3) | 191 (24.2) | 0.021 |
| **Hypertension (%)** |  | 1927 (74.4) | 579 (73.3) | 0.025 | 563 (71.3) | 579 (73.3) | 0.045 |
| **Substance Use Disorder (%)** |  | 862 (33.3) | 245 (31.0) | 0.049 | 230 (29.1) | 245 (31.0) | 0.041 |
| **Depression (%)** |  | 1094 (42.2) | 487 (61.6) | 0.396 | 483 (61.1) | 487 (61.6) | 0.010 |
| **Hypertension Medication (%)** |  | 1615 (62.4) | 487 (61.6) | 0.015 | 489 (61.9) | 487 (61.6) | 0.005 |
| **T2DM Medication (%)** |  | 353 (13.6) | 130 (16.5) | 0.079 | 130 (16.5) | 130 (16.5) | <0.001 |
| **Lipid Lowering Medication (%)** |  | 1182 (45.6) | 389 (49.2) | 0.072 | 370 (46.8) | 389 (49.2) | 0.048 |
| **Benzodiazepines Medication (%)** |  | 12 (0.5) | 1 (0.1) | 0.062 | 0 (0.0) | 1 (0.1) | 0.050 |
| **Antidepressant Medication (%)** |  | 1431 (55.3) | 551 (69.7) | 0.303 | 558 (70.6) | 551 (69.7) | 0.019 |
| **AD Medication (%)** |  | 1277 (49.3) | 417 (52.8) | 0.070 | 423 (53.5) | 417 (52.8) | 0.015 |

|  |  |  |  |  |  |  |
| --- | --- | --- | --- | --- | --- | --- |
| **Anticholinergics Medication (%)** | 140 (5.4) | 62 (7.8) | 0.098 | 65 (8.2) | 62 (7.8) | 0.014 |
| **Analgesics Medication (%)** | 256 (9.9) | 76 (9.6) | 0.009 | 63 (8.0) | 76 (9.6) | 0.058 |

Abbreviations: SMD, standardized mean difference; SD, standard deviation; T2DM, Type 2 Diabetes Mellitus; AD, Alzheimer's Disease

### Comparative All-Cause Mortality Risk

Time-varying Cox proportional hazards models were used to estimate the comparative risk of all-cause mortality among the six drug comparison pairs. The pooled results are summarized in **Figure 1**. Kaplan–Meier survival curves were generated for descriptive purposes and presented in the supplementary materials (**Supplementary Figure 1-6**). Aripiprazole was associated with a significantly lower risk of mortality when compared to Olanzapine (AHR = 0.667, 95% CI 0.472–0.941, p = 0.021) and Quetiapine (AHR = 0.677, 95% CI 0.462–0.990, p = 0.044).

**Figure 1.**
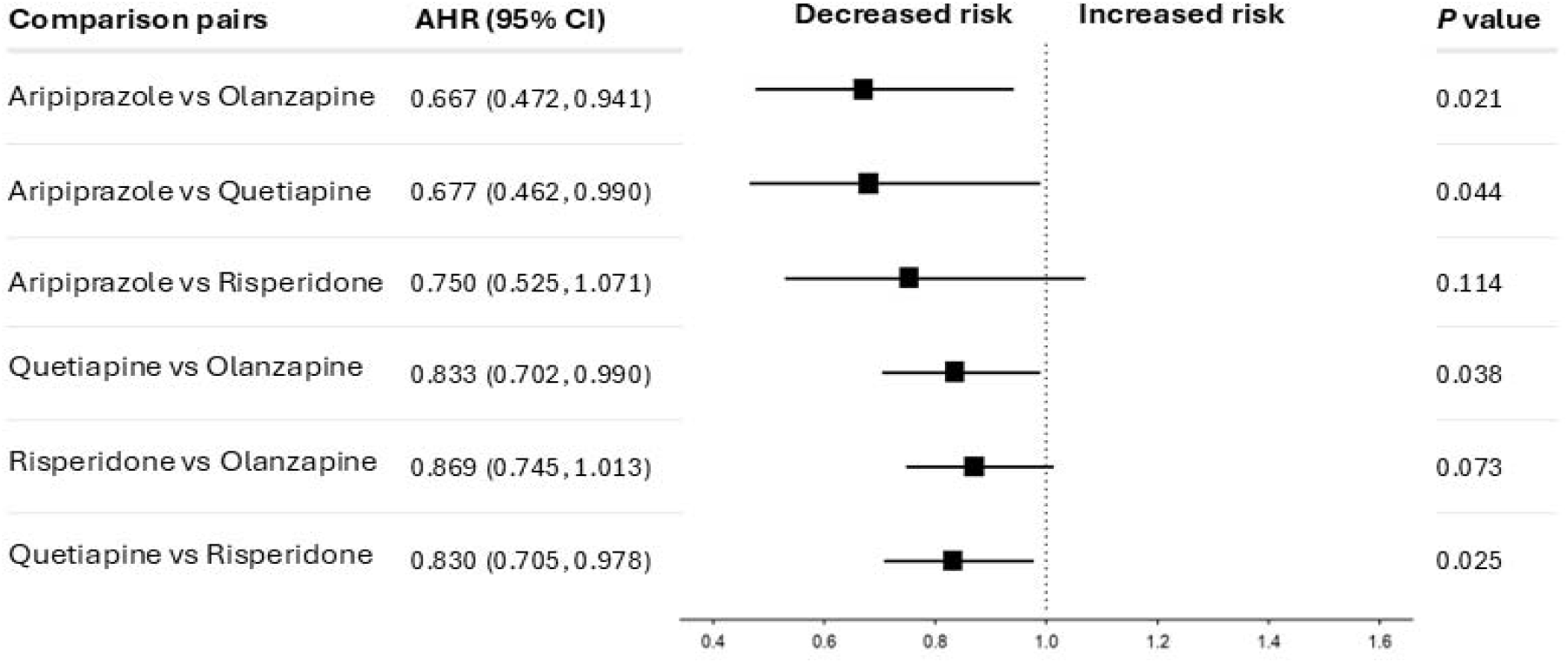
Adjusted Hazard Ratios (AHRs) for All-Cause Mortality for Time-Varying Cox Proportional Hazards Regression Models.

Additionally, Quetiapine was associated with a significantly lower risk of mortality when compared to Olanzapine (AHR = 0.833, 95% CI 0.702–0.990, p = 0.038) and Risperidone (AHR = 0.830, 95% CI 0.705–0.978, p = 0.026).

### Sensitivity Analysis

Multiple sensitivity analyses were conducted by altering the grace period definitions (14, 60, and 90 days) and restricting follow-up to one year (shown in **Supplementary Table S6**). Specifically, the protective effect of aripiprazole versus olanzapine remained statistically significant across all sensitivity analyses, with hazard ratios ranging from 0.583 to 0.741. Quetiapine consistently demonstrated lower mortality risk compared to Olanzapine, with HRs between 0.773 and 0.833 across all scenarios. However, some estimates showed sensitivity to parameter changes. For instance, the mortality benefit of aripiprazole over quetiapine lost significance under a 14-day grace period (HR = 0.800, p = 0.296) and under a 1-year follow-up window (HR = 0.697, p = 0.064), although HR value remained similar. In contrast, the previously non-significant comparison between aripiprazole and risperidone became statistically significant when the grace period was extended to 90 days (HR = 0.677, p = 0.0045). The significance of quetiapine versus risperidone also varied, disappearing under 14-day,60-day and 90-day windows, but re-emerging with a 1-year follow-up period (HR = 0.841, p = 0.0378).

### Treatment Effect Heterogeneity

To explore variation in treatment effects across patient subgroups, causal tree models were used for each of the six drug comparison pairs. This model divided the study population into mutually exclusive subgroups with different treatment responses, as quantified by the average treatment effect (ATE), which was defined as the absolute difference in the probability of death within the compared medications. Subgroups with statistically significant ATE are shown in **Figure 2, 3 and Supplementary Figure 7-10**. Notably, use of T2DM medications and marital status frequently emerged as important effect modifiers across multiple comparisons.

**Figure 2.**
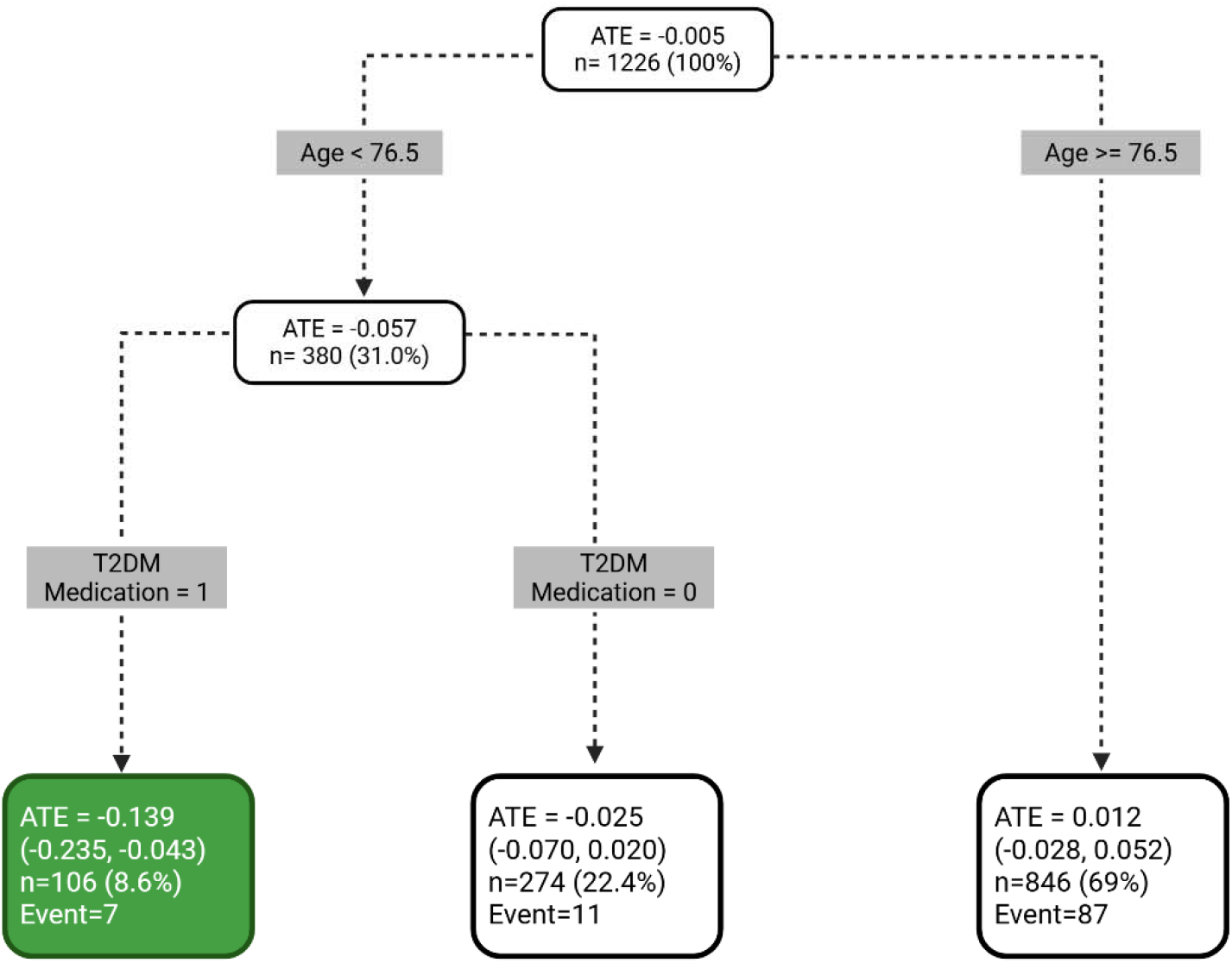
Subgroups with heterogeneous treatment effects on all-cause mortality among aripiprazole vs. quetiapine. Causal tree analysis identified patient subgroups with significantly different average treatment effects (ATEs) of aripiprazole compared to quetiapine. Terminal nodes represent subgroups, with corresponding ATE values. Negative ATEs indicate reduced mortality risk associated with aripiprazole. All estimates were derived from the matched analytic sample. Binary covariates (e.g., T2DM Medication; Depression) are coded as 1 = presence/yes and 0 = absence/no.

**Figure 3.**
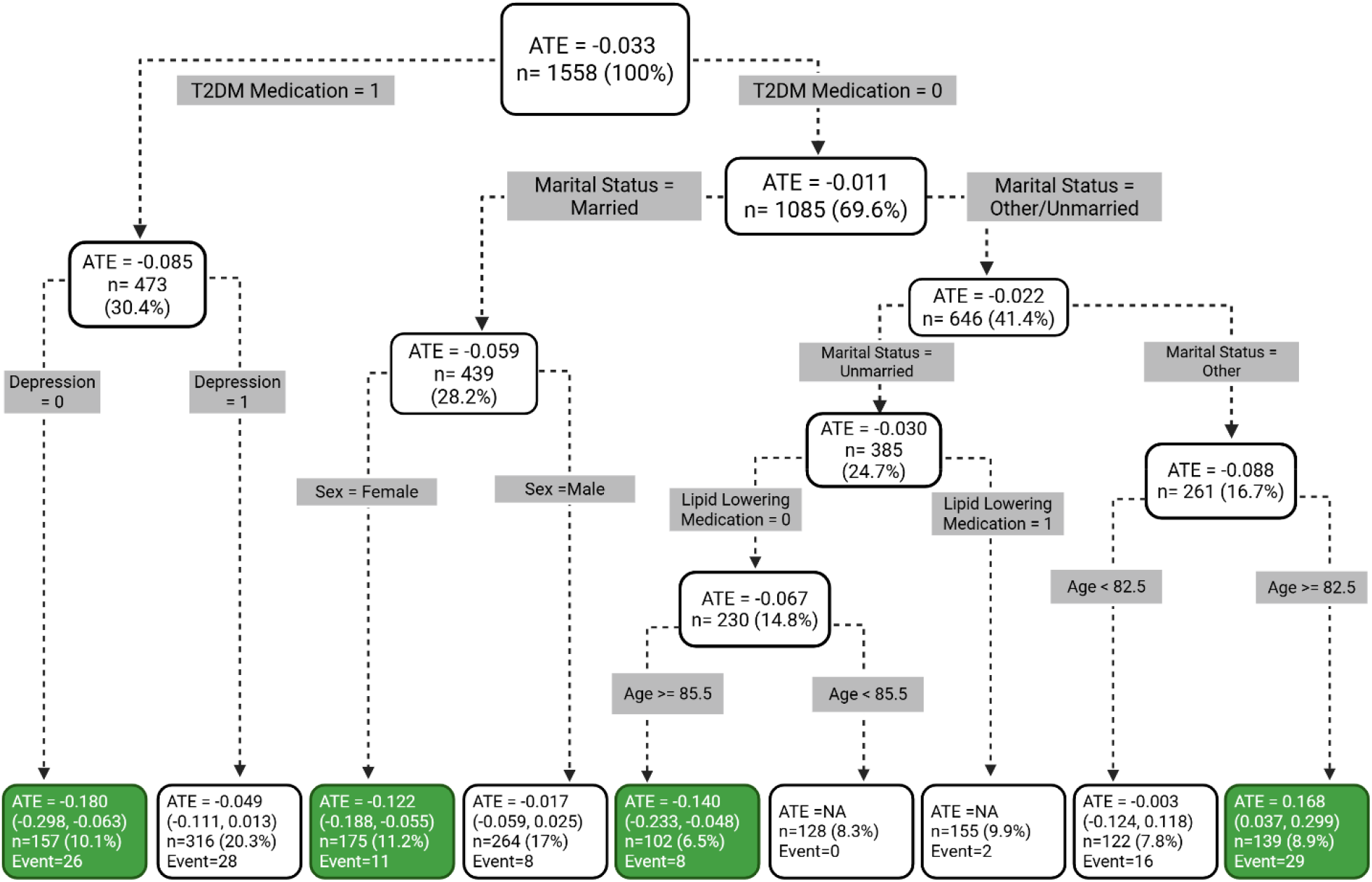
Subgroups with heterogeneous treatment effects on all-cause mortality among aripiprazole vs. risperidone. Causal tree analysis identified patient subgroups with significantly different average treatment effects (ATEs) of aripiprazole compared to risperidone. Terminal nodes represent subgroups, with corresponding ATE values. Negative ATEs indicate reduced mortality risk associated with aripiprazole. All estimates were derived from the matched analytic sample. Binary covariates (e.g., T2DM Medication; Depression) are coded as 1 = presence/yes and 0 = absence/no.

In the comparison between aripiprazole and olanzapine, stronger protective effects of aripiprazole were observed among patients with cancer who were also using lipid-lowering medications (ATE = -0.135; 95% CI, -0.226 ∼ - 0.045), and among females concurrently using anti-dementia medications (ATE = -0.108; 95% CI, -0.19 ∼ -0.026). When comparing aripiprazole to quetiapine, patients younger than 76.5 years and receiving T2DM medications exhibited a significantly larger benefit from aripiprazole (ATE = -0.139; 95% CI, -0.235 ∼ -0.043). A similar pattern was found in the aripiprazole versus risperidone analysis, where patients with T2DM but without depression showed the most pronounced treatment effect (ATE = -0.180; 95% CI, -0.298 ∼ -0.063).

In the olanzapine versus quetiapine comparison, treatment effect heterogeneity was evident among patients concurrently using antihypertensive and anti-dementia medications (ATE = 0.086; 95% CI, 0.023 ∼ 0.148), as well as among those aged 82.5 years or older (ATE = 0.035; 95% CI, 0.002 ∼ 0.068). The comparison between olanzapine and risperidone identified a high-risk subgroup consisting of patients without substance use disorder (SUD), using antidepressants, and with no history of depression (ATE = -0.169; 95% CI, -0.267 ∼ -0.071). Finally, in the risperidone versus quetiapine analysis, patients without heart failure but with SUD and aged ≥85.5 benefited more from quetiapine (ATE = -0.130; 95% CI, -0.260 ∼ -0.0001), whereas patients with heart failure aged between 82.5 and 91.5 years showed greater benefit from risperidone (ATE = 0.201; 95% CI, 0.107 ∼ 0.295).

### Subgroup Cox Analyses Based on Heterogeneity Findings

Given that T2DM medication use emerged as a potential modifier of treatment effects across 2 comparisons in the causal tree models, we conducted additional subgroup Cox analyses to further assess its impact. In the subgroup of patients using T2DM medications, aripiprazole showed a trend towards reduced mortality risk compared to risperidone (HR = 0.375, 95% CI: 0.134–1.051, p = 0.062), and significantly lower risk when compared to the combined group of risperidone and quetiapine (HR = 0.604, 95% CI: 0.439–0.831, p = 0.002). In contrast, similar analyses within the subgroup of patients using anti-dementia medications did not yield statistically significant results. These results are shown in **Supplementary Table S7**.

## Discussion

In this large real-world cohort of patients with Alzheimer’s disease (AD), we observed notable differences in all-cause mortality risk across several commonly prescribed SGAs. Using time-varying Cox models, we identified significant differences in mortality risk between different SGAs treatment, with aripiprazole and quetiapine showing lower associated mortality in several head-to-head comparisons. To explore potential treatment effect heterogeneity, we further applied causal tree models to identify subgroups with differential treatment responses. Across multiple comparisons, factors such as concurrent use of anti-dementia or T2DM medications appeared to influence the magnitude of treatment effects. These findings provide strong evidence on the necessary personalized risk-benefit evaluation when prescribing antipsychotics in AD populations.

Our survival analysis revealed that different SGAs are associated with distinct risks of all-cause mortality in patients with AD. Many prior studies have demonstrated that antipsychotic use, particularly among older adults with dementia, is linked to increased mortality[19-21]. This evidence led to the FDA’s black box warning in 2005 regarding the use of these medications in dementia-related psychosis[4]. However, these existing studies have focused on comparisons between overall antipsychotics and placebo or non-user. In contrast, our study used an active-comparator design to directly compare single SGAs within a well-matched AD population, our findings are also supported by previous studies reporting similar safety patterns among SGAs. Kales et al. reported that, among patients with dementia in the U.S. VA system, quetiapine was associated with the lowest mortality risk when compared to risperidone, and there is no significant difference in mortality between olanzapine and risperidone[22]. This pattern aligns with our findings. However, this study was based on a predominantly male veteran population, which limits its generalizability. Our data source had a more balanced gender distribution, potentially improving the external validity of our results. Similarly, Rossom et al. observed differential mortality risks when individual antipsychotics were compared to no treatment, with quetiapine showing the lowest hazard ratio, followed by olanzapine and risperidone [23]. While this study also suggests a safety hierarchy among SGAs, the comparison between treatment users and non-users raises concerns of indication bias. For our study, under a real-world clinical decision-making setting, we care more about which specific antipsychotic poses the least risk when treatment is necessary. Real world evidence for head-to-head comparisons involving aripiprazole remains limited. Two network meta-analyses included aripiprazole but differed from our study in several prospectives. Yunusa et al. found no statistically significant differences in mortality among SGAs, but their conclusions were based on indirect comparisons, and the inclusion criteria were restricted to patients with dementia-related psychosis rather than an AD population[24]. Lü et al. did not examine aripiprazole’s impact on mortality, but rather emphasized its relative advantage in terms of acceptability, which limits the comparability of its findings to our results[25].

The consistently lower hazard ratios observed for aripiprazole in our study may be partly attributable to its favorable metabolic profile. Compared to other SGAs, aripiprazole exhibits a relatively favorable metabolic profile, with lower incidences of weight gain, glucose dysregulation, and dyslipidemia[26]. These properties may collectively reduce cardiovascular and metabolic complications, which are key contributors to mortality in patients with AD[27]. This explanation is further supported by our subgroup analysis, which found that the mortality benefit of aripiprazole was more notable among patients concurrently using type 2 diabetes medications. Additionally, unlike olanzapine, risperidone, and quetiapine, aripiprazole is a partial agonist at D□and 5-HT□A receptors, which allows it to modulate dopaminergic activity in a more balanced manner rather than fully blocking the receptor[28]. This mechanism has been associated with a lower risk of extrapyramidal symptoms (EPS), which are known to contribute to morbidity and mortality in older adults[28, 29].

Building on these overall mortality differences across SGAs, we further explored whether certain patient subgroups might experience differential benefits from specific medications. Using causal tree analysis with TMLE, we identified treatment effect heterogeneity (HTE) based on baseline characteristics. In particular, among AD patients who were using T2DM medications, which is an indicator of pre□existing metabolic dysregulation, we observed that the survival benefit of Aripiprazole over Quetiapine (ATE ≈□−0.139) and over Risperidone (ATE ≈□−0.180) was more obvious than in the overall cohort. This finding may be explained by existing evidence that SGAs differ substantially in their metabolic impact. Citrome et al. reported that aripiprazole has been shown to carry a relatively low risk of inducing weight gain, insulin resistance and new□onset T2DM in adult populations[30]. Although Yeh et al. observed a slightly increased risk of long-term major adverse cardiovascular events with aripiprazole compared to risperidone in patients with schizophrenia and T2DM, the risk of sudden cardiac death was numerically lower for aripiprazole in both the first year and beyond one year[31]. While the patient populations and outcome definitions are different from ours, this trend aligns with the mortality benefit observed in our AD cohort.

Sensitivity analyses demonstrated that the main findings were generally robust to variations in grace period definitions and follow-up periods. While most hazard ratios maintained similar directions and magnitudes across different grace periods (14, 60, and 90 days) and a shorter 1-year follow-up, statistical significance varied in a few comparisons. The comparison between risperidone and quetiapine demonstrated statistical significance only under the 30-day grace period and became marginally significant under the 90-day window. This suggests that the observed treatment effect for this comparison may be sensitive to exposure risk window definition. Moreover, when a shorter 14-day grace period was applied, several comparison pairs lost statistical significance, which suggests that the mortality risks associated with these antipsychotic medications may not emerge immediately after treatment ends. Instead, their effects on mortality may develop gradually over a longer duration. These findings showed the methodological importance of defining an appropriate grace period in pharmacoepidemiologic studies, as it can substantially impact the interpretation of treatment effects.

This study has several strengths. We used real-world electronic health record data from a large and diverse Alzheimer’s disease population. Importantly, the Truveta platform includes linked pharmacy dispensing data, which enhances the accuracy of medication exposure. We applied active-comparator, new-user design and time-varying Cox models were applied, which could improve upon traditional designs by reducing indication bias and better reflecting clinical practice. Given the high prevalence of medication non-adherence and discontinuation in real-world settings, modeling drug exposure as a time-varying variable allowed us to more accurately capture patients’ dynamic treatment patterns. Furthermore, we applied a causal machine learning framework, causal tree analysis with targeted maximum likelihood estimation (TMLE), to explore treatment effect heterogeneity. This method enabled data-driven identification of clinically meaningful subgroups that may benefit differently from specific SGAs, providing insights for future work in personalized prescribing.

However, several limitations should be noted. Our data did not include detailed clinical assessments of neuropsychiatric symptom severity, which could influence both treatment decisions and outcomes. While drug exposure was defined using dispense records, actual medication adherence could not be confirmed. Additionally, we focused on all-cause mortality rather than cause-specific death. Future research using more detailed mortality data may help explore the specific pathways through which SGAs influence mortality in this population. Survival times observed in our Kaplan-Meier curves may appear higher than those reported in earlier prospective studies [32]. This may reflect differences in follow-up structure, as Truveta may not capture patients who later transition to nonparticipating systems, leading to right censoring. To minimize the bias and ensure robust estimates, we limited follow-up to two years, during which our observed survival patterns were consistent with recent population-level estimates[33]. Lastly, although the overall sample was large, some subgroups identified through causal tree analysis were relatively small, and future studies with more eligible participants are needed to validate these findings.

## Conclusion

In this real-world cohort of Alzheimer’s disease patients, we found significant differences in all-cause mortality across commonly prescribed second-generation antipsychotics. Aripiprazole was associated with lower mortality compared to olanzapine and quetiapine, while quetiapine showed lower mortality than olanzapine and risperidone. Additionally, causal tree analysis revealed that certain clinical characteristics, such as use of T2DM medications, may modify the relative mortality benefits of specific SGAs, highlighting the importance of individualized prescribing in this population.

## Data Availability

The data used in this study are not publicly available but may be obtained from Truveta under a data use agreement. Results and code are available from the authors upon reasonable request.

## References

1. Aarsland D. Epidemiology and Pathophysiology of Dementia-Related Psychosis. J Clin Psychiatry. 2020 Sep 15;81(5).

2. Alexander GC, Gallagher SA, Mascola A, Moloney RM, Stafford RS. Increasing off-label use of antipsychotic medications in the United States, 1995-2008. Pharmacoepidemiol Drug Saf. 2011 Feb;20(2):177–84.

3. Rogowska M, Thornton M, Creese B, Velayudhan L, Aarsland D, Ballard C, et al. Implications of Adverse Outcomes Associated with Antipsychotics in Older Patients with Dementia: A 2011-2022 Update. Drugs Aging. 2023 Jan;40(1):21–32.

4. Administration FaD. Public health advisory: deaths with antipsychotics in elderly patients with behavioral disturbances. 2005.

5. Mok PLH, Carr MJ, Guthrie B, Morales DR, Sheikh A, Elliott RA, et al. Multiple adverse outcomes associated with antipsychotic use in people with dementia: population based matched cohort study. BMJ. 2024;385:e076268.

6. Ralph SJ, Espinet AJ. Increased All-Cause Mortality by Antipsychotic Drugs: Updated Review and Meta-Analysis in Dementia and General Mental Health Care. Journal of Alzheimer’s Disease Reports. 2018;2(1):1–26.

7. Santiago JA, Potashkin JA. The Impact of Disease Comorbidities in Alzheimer’s Disease. Front Aging Neurosci. 2021;13:631770.

8. von Elm E, Altman DG, Egger M, Pocock SJ, Gøtzsche PC, Vandenbroucke JP. The Strengthening the Reporting of Observational Studies in Epidemiology (STROBE) statement: guidelines for reporting observational studies. Lancet. 2007 Oct 20;370(9596):1453–7.

9. Bharat C, Degenhardt L, Pearson SA, Buizen L, Wilson A, Dobbins T, et al. A data-informed approach using individualised dispensing patterns to estimate medicine exposure periods and dose from pharmaceutical claims data. Pharmacoepidemiol Drug Saf. 2023 Mar;32(3):352–65.

10. Filion KB, Lix LM, Yu OH, Dell’Aniello S, Douros A, Shah BR, et al. Sodium glucose cotransporter 2 inhibitors and risk of major adverse cardiovascular events: multi-database retrospective cohort study. Bmj. 2020 Sep 23;370:m3342.

11. Bjelkarøy MT, Simonsen TB, Siddiqui Tahreem Ghazal, Cheng S, Grambaite R, Benth JŠ, et al. Mortality and health-related quality of life in older adults with long-term use of opioids, z-hypnotics or benzodiazepines: a prospective observational study at 5 years follow-up. BMJ Open. 2024;14(2):e079347.

12. Xu KY, Hartz SM, Borodovsky JT, Bierut LJ, Grucza RA. Association Between Benzodiazepine Use With or Without Opioid Use and All-Cause Mortality in the United States, 1999-2015. JAMA Network Open. 2020;3(12):e2028557–e.

13. Ali S, Peterson GM, Bereznicki LR, Salahudeen MS. Association between anticholinergic drug burden and mortality in older people: a systematic review. European Journal of Clinical Pharmacology. 2020 2020/03/01;76(3):319–35.

14. Gislason GH, Rasmussen JN, Abildstrom SZ, Schramm TK, Hansen ML, Fosbøl EL, et al. Increased Mortality and Cardiovascular Morbidity Associated With Use of Nonsteroidal Anti-inflammatory Drugs in Chronic Heart Failure. Archives of Internal Medicine. 2009;169(2):141–9.

15. Ho D, Imai K, King G, Stuart EA. MatchIt: Nonparametric Preprocessing for Parametric Causal Inference. Journal of Statistical Software. 2011 06/14;42(8):1–28.

16. Wager S, Athey S. Estimation and Inference of Heterogeneous Treatment Effects using Random Forests. Journal of the American Statistical Association. 2018 2018/07/03;113(523):1228–42.

17. Gruber S, Van Der Laan M. tmle: an R package for targeted maximum likelihood estimation. Journal of Statistical Software. 2012;51:1–35.

18. van der Laan MJ, Polley EC, Hubbard AE. Super learner. Stat Appl Genet Mol Biol. 2007;6:Article25.

19. Gardette V, Lapeyre-Mestre M, Coley N, Cantet C, Montastruc JL, Vellas B, et al. Antipsychotic use and mortality risk in community-dwelling Alzheimer’s disease patients: evidence for a role of dementia severity. Curr Alzheimer Res. 2012 Nov;9(9):1106–16.

20. Jennum P, Baandrup L, Ibsen R, Kjellberg J. Increased all-cause mortality with use of psychotropic medication in dementia patients and controls: A population-based register study. Eur Neuropsychopharmacol. 2015 Nov;25(11):1906–13.

21. Nielsen RE, Lolk A, Valentin JB, Andersen K. Cumulative dosages of antipsychotic drugs are associated with increased mortality rate in patients with Alzheimer’s dementia. Acta Psychiatr Scand. 2016 Oct;134(4):314–20.

22. Kales HC, Kim HM, Zivin K, Valenstein M, Seyfried LS, Chiang C, et al. Risk of mortality among individual antipsychotics in patients with dementia. Am J Psychiatry. 2012 Jan;169(1):71–9.

23. Rossom RC, Rector TS, Lederle FA, Dysken MW. Are all commonly prescribed antipsychotics associated with greater mortality in elderly male veterans with dementia? J Am Geriatr Soc. 2010 Jun;58(6):1027–34.

24. Yunusa I, Rashid N, Demos GN, Mahadik BS, Abler VC, Rajagopalan K. Comparative Outcomes of Commonly Used Off-Label Atypical Antipsychotics in the Treatment of Dementia-Related Psychosis: A Network Meta-analysis. Adv Ther. 2022 May;39(5):1993–2008.

25. Lü W, Liu F, Zhang Y, He X, Hu Y, Xu H, et al. Efficacy, acceptability and tolerability of second-generation antipsychotics for behavioural and psychological symptoms of dementia: a systematic review and network meta-analysis. BMJ Ment Health. 2024 Jul 30;27(1).

26. Scigliano G, Ronchetti G. Antipsychotic-induced metabolic and cardiovascular side effects in schizophrenia: a novel mechanistic hypothesis. CNS Drugs. 2013 Apr;27(4):249–57.

27. Liu L, Gracely EJ, Zhao X, Gliebus GP, May NS, Volpe SL, et al. Association of multiple metabolic and cardiovascular markers with the risk of cognitive decline and mortality in adults with Alzheimer’s disease and AD-related dementia or cognitive decline: a prospective cohort study. Front Aging Neurosci. 2024;16:1361772.

28. Gettu N SA. Aripiprazole. 2023 May 16 [cited; Available from: https://www.ncbi.nlm.nih.gov/books/NBK547739/

29. Kadakia A, Brady BL, Dembek C, Williams GR, Kent JM. The incidence and economic burden of extrapyramidal symptoms in patients with schizophrenia treated with second generation antipsychotics in a Medicaid population. J Med Econ. 2022 Jan-Dec;25(1):87–98.

30. Citrome L, Kalsekar I, Baker RA, Hebden T. A review of real-world data on the effects of aripiprazole on weight and metabolic outcomes in adults. Curr Med Res Opin. 2014 Aug;30(8):1629–41.

31. Yeh YT, Lo SC, Huang CN, Yang YS, Liao PL, Kornelius E. Comparative cardiovascular outcomes of aripiprazole vs. risperidone in patients with type 2 diabetes and schizophrenia: a retrospective cohort study. Front Pharmacol. 2025;16:1617534.

32. Brookmeyer R, Corrada MM, Curriero FC, Kawas C. Survival Following a Diagnosis of Alzheimer Disease. Archives of Neurology. 2002;59(11):1764–7.

33. Brück CC, Mooldijk SS, Kuiper LM, Sambou ML, Licher S, Mattace-Raso F, et al. Time to nursing home admission and death in people with dementia: systematic review and meta-analysis. BMJ. 2025;388:e080636.

