## Supplementary Figure 1-10; Supplementary Table1-7 for "Comparative Mortality Risk of Aripiprazole, Olanzapine, Quetiapine and Risperidone in Alzheimer’s Disease: A Real□World Cohort Study with Treatment Effect Heterogeneity Analysis"

**Supplementary Table 1 Baseline Characteristics of Patients Treated with Aripiprazole and Quetiapine Before and After Propensity Score Matching**

|  |  | **Before Propensity Score Matching** | | | **After Propensity Score Matching** | | |
| --- | --- | --- | --- | --- | --- | --- | --- |
|  |  | **Quetiapine** | **Aripiprazole** | **SMD** | **Quetiapine** | **Aripiprazole** | **SMD** |
| **n** |  | 10938 | 613 |  | 613 | 613 |  |
| **Age (SD)** |  | 84.31 (7.98) | 79.91 (9.42) | 0.504 | 79.51 (9.06) | 79.91 (9.42) | 0.044 |
| **Sex (%)** | Female | 6696 (61.2) | 435 (71.0) | 0.207 | 441 (71.9) | 435 (71.0) | 0.022 |
|  | Male | 4242 (38.8) | 178 (29.0) |  | 172 (28.1) | 178 (29.0) |  |
| **Race (%)** | African American | 741 (6.8) | 31 (5.1) | 0.082 | 25 (4.1) | 31 (5.1) | 0.066 |
|  | Other | 483 (4.4) | 23 (3.8) |  | 29 (4.7) | 23 (3.8) |  |
|  | White | 9714 (88.8) | 559 (91.2) |  | 559 (91.2) | 559 (91.2) |  |
| **Marital Status (%)** | Married | 3896 (35.6) | 236 (38.5) | 0.159 | 241 (39.3) | 236 (38.5) | 0.028 |
|  | Other/Unknown | 3773 (34.5) | 167 (27.2) |  | 170 (27.7) | 167 (27.2) |  |
|  | Unmarried | 3269 (29.9) | 210 (34.3) |  | 202 (33.0) | 210 (34.3) |  |
| **Heart Failure (%)** | | 1963 (17.9) | 99 (16.2) | 0.048 | 90 (14.7) | 99 (16.2) | 0.041 |
| **Myocardial Infarction (%)** | | 1036 (9.5) | 40 (6.5) | 0.109 | 30 (4.9) | 40 (6.5) | 0.070 |
| **Stroke (%)** | | 1789 (16.4) | 89 (14.5) | 0.051 | 72 (11.7) | 89 (14.5) | 0.082 |
| **Chronic Kidney Disease (%)** | | 3205 (29.3) | 181 (29.5) | 0.005 | 164 (26.8) | 181 (29.5) | 0.062 |
| **Chronic Obstructive Pulmonary Disease (%)** | | 1486 (13.6) | 109 (17.8) | 0.116 | 90 (14.7) | 109 (17.8) | 0.084 |
| **Type 2 Diabetes Mellitus (%)** | | 3002 (27.4) | 186 (30.3) | 0.064 | 169 (27.6) | 186 (30.3) | 0.061 |
| **Cancer (%)** | | 2983 (27.3) | 146 (23.8) | 0.079 | 149 (24.3) | 146 (23.8) | 0.011 |
| **Hypertension (%)** | | 8435 (77.1) | 451 (73.6) | 0.082 | 430 (70.1) | 451 (73.6) | 0.076 |
| **Substance Use Disorder (%)** | | 3190 (29.2) | 192 (31.3) | 0.047 | 191 (31.2) | 192 (31.3) | 0.004 |
| **Depression (%)** | | 4165 (38.1) | 398 (64.9) | 0.558 | 391 (63.8) | 398 (64.9) | 0.024 |
| **Hypertension Medication (%)** | | 6840 (62.5) | 385 (62.8) | 0.006 | 365 (59.5) | 385 (62.8) | 0.067 |
| **T2DM Medication (%)** | | 1425 (13.0) | 103 (16.8) | 0.106 | 93 (15.2) | 103 (16.8) | 0.045 |
| **Lipid Lowering Medication (%)** | | 5034 (46.0) | 307 (50.1) | 0.081 | 288 (47.0) | 307 (50.1) | 0.062 |
| **Benzodiazepines Medication (%)** | | 40 (0.4) | 1 (0.2) | 0.039 | 2 (0.3) | 1 (0.2) | 0.033 |
| **Antidepressant Medication (%)** | | 5814 (53.2) | 449 (73.2) | 0.426 | 441 (71.9) | 449 (73.2) | 0.029 |
| **AD Medication (%)** | | 5780 (52.8) | 334 (54.5) | 0.033 | 341 (55.6) | 334 (54.5) | 0.023 |
| **Anticholinergics Medication (%)** | | 656 (6.0) | 47 (7.7) | 0.066 | 38 (6.2) | 47 (7.7) | 0.058 |
| **Analgesics Medication (%)** | | 956 (8.7) | 60 (9.8) | 0.036 | 45 (7.3) | 60 (9.8) | 0.088 |

Abbreviations: SMD, standardized mean difference; SD, standard deviation; T2DM, Type 2 Diabetes Mellitus; AD, Alzheimer’s Disease

|  |  | **Before Propensity Score Matching** | | | **After Propensity Score Matching** | | |
| --- | --- | --- | --- | --- | --- | --- | --- |
|  |  | **Risperidone** | **Aripiprazole** | **SMD** | **Risperidone** | **Aripiprazole** | **SMD** |
| **n** |  | 2910 | 779 |  | 779 | 779 |  |
| **Age (SD)** |  | 83.67 (8.28) | 80.01 (9.37) | 0.414 | 80.28 (9.07) | 80.01 (9.37) | 0.029 |
| **Sex (%)** | Female | 1855 (63.7) | 545 (70.0) | 0.132 | 549 (70.5) | 545 (70.0) | 0.011 |
|  | Male | 1055 (36.3) | 234 (30.0) |  | 230 (29.5) | 234 (30.0) |  |
| **Race (%)** | African American | 205 (7.0) | 40 (5.1) | 0.083 | 43 (5.5) | 40 (5.1) | 0.037 |
|  | Other | 123 (4.2) | 30 (3.9) |  | 35 (4.5) | 30 (3.9) |  |
|  | White | 2582 (88.7) | 709 (91.0) |  | 701 (90.0) | 709 (91.0) |  |
| **Marital Status (%)** | Married | 974 (33.5) | 301 (38.6) | 0.184 | 301 (38.6) | 301 (38.6) | 0.007 |
|  | Other/Unknown | 1018 (35.0) | 207 (26.6) |  | 205 (26.3) | 207 (26.6) |  |
|  | Unmarried | 918 (31.5) | 271 (34.8) |  | 273 (35.0) | 271 (34.8) |  |
| **Heart Failure (%)** | | 479 (16.5) | 125 (16.0) | 0.011 | 120 (15.4) | 125 (16.0) | 0.018 |
| **Myocardial Infarction (%)** | | 246 (8.5) | 58 (7.4) | 0.037 | 59 (7.6) | 58 (7.4) | 0.005 |
| **Stroke (%)** | | 435 (14.9) | 123 (15.8) | 0.023 | 128 (16.4) | 123 (15.8) | 0.017 |
| **Chronic Kidney Disease (%)** | | 796 (27.4) | 231 (29.7) | 0.051 | 235 (30.2) | 231 (29.7) | 0.011 |
| **Chronic Obstructive Pulmonary Disease (%)** | | 398 (13.7) | 128 (16.4) | 0.077 | 141 (18.1) | 128 (16.4) | 0.044 |
| **Type 2 Diabetes Mellitus (%)** | | 734 (25.2) | 237 (30.4) | 0.116 | 236 (30.3) | 237 (30.4) | 0.003 |
| **Cancer (%)** | | 739 (25.4) | 192 (24.6) | 0.017 | 193 (24.8) | 192 (24.6) | 0.003 |
| **Hypertension (%)** | | 2170 (74.6) | 574 (73.7) | 0.020 | 585 (75.1) | 574 (73.7) | 0.032 |
| **Substance Use Disorder (%)** | | 838 (28.8) | 251 (32.2) | 0.074 | 246 (31.6) | 251 (32.2) | 0.014 |
| **Depression (%)** | | 1171 (40.2) | 480 (61.6) | 0.438 | 487 (62.5) | 480 (61.6) | 0.019 |
| **Hypertension Medication (%)** | | 1746 (60.0) | 486 (62.4) | 0.049 | 494 (63.4) | 486 (62.4) | 0.021 |
| **T2DM Medication (%)** | | 365 (12.5) | 130 (16.7) | 0.118 | 123 (15.8) | 130 (16.7) | 0.024 |
| **Lipid Lowering Medication (%)** | | 1260 (43.3) | 386 (49.6) | 0.126 | 396 (50.8) | 386 (49.6) | 0.026 |
| **Benzodiazepines Medication (%)** | | 9 (0.3) | 3 (0.4) | 0.013 | 5 (0.6) | 3 (0.4) | 0.036 |
| **Antidepressant Medication (%)** | | 1632 (56.1) | 553 (71.0) | 0.313 | 551 (70.7) | 553 (71.0) | 0.006 |
| **AD Medication (%)** | | 1563 (53.7) | 408 (52.4) | 0.027 | 424 (54.4) | 408 (52.4) | 0.041 |
| **Anticholinergics Medication (%)** | | 149 (5.1) | 57 (7.3) | 0.091 | 49 (6.3) | 57 (7.3) | 0.041 |
| **Analgesics Medication (%)** | | 234 (8.0) | 75 (9.6) | 0.056 | 66 (8.5) | 75 (9.6) | 0.040 |

**Supplementary Table 2 Baseline Characteristics of Patients Treated with Aripiprazole and Risperidone Before and After Propensity Score Matching**

Abbreviations: SMD, standardized mean difference; SD, standard deviation; T2DM, Type 2 Diabetes Mellitus; AD, Alzheimer’s Disease

**Supplementary Table 3 Baseline Characteristics of Patients Treated with Olanzapine and Quetiapine Before and After Propensity Score Matching**

|  |  | **Before Propensity Score Matching** | | | **After Propensity Score Matching** | | |
| --- | --- | --- | --- | --- | --- | --- | --- |
|  |  | **Quetiapine** | **Olanzapine** | **SMD** | **Quetiapine** | **Olanzapine** | **SMD** |
| **n** |  | 10113 | 1609 |  | 1609 | 1609 |  |
| **Age (SD)** |  | 84.41 (7.98) | 83.25 (8.65) | 0.139 | 83.11 (8.44) | 83.25 (8.65) | 0.016 |
| **Sex (%)** | Female | 6224 (61.5) | 1015 (63.1) | 0.032 | 1035 (64.3) | 1015 (63.1) | 0.026 |
|  | Male | 3889 (38.5) | 594 (36.9) |  | 574 (35.7) | 594 (36.9) |  |
| **Race (%)** | African American | 695 (6.9) | 100 (6.2) | 0.052 | 88 (5.5) | 100 (6.2) | 0.034 |
|  | Other | 450 (4.4) | 58 (3.6) |  | 55 (3.4) | 58 (3.6) |  |
|  | White | 8968 (88.7) | 1451 (90.2) |  | 1466 (91.1) | 1451 (90.2) |  |
| **Marital Status (%)** | Married | 3591 (35.5) | 519 (32.3) | 0.072 | 525 (32.6) | 519 (32.3) | 0.043 |
|  | Other/Unknown | 3459 (34.2) | 564 (35.1) |  | 533 (33.1) | 564 (35.1) |  |
|  | Unmarried | 3063 (30.3) | 526 (32.7) |  | 551 (34.2) | 526 (32.7) |  |
| **Heart Failure (%)** | | 1837 (18.2) | 306 (19.0) | 0.022 | 307 (19.1) | 306 (19.0) | 0.002 |
| **Myocardial Infarction (%)** | | 965 (9.5) | 164 (10.2) | 0.022 | 177 (11.0) | 164 (10.2) | 0.026 |
| **Stroke (%)** | | 1658 (16.4) | 272 (16.9) | 0.014 | 260 (16.2) | 272 (16.9) | 0.020 |
| **Chronic Kidney Disease (%)** | | 2973 (29.4) | 466 (29.0) | 0.010 | 481 (29.9) | 466 (29.0) | 0.020 |
| **Chronic Obstructive Pulmonary Disease (%)** | | 1382 (13.7) | 263 (16.3) | 0.075 | 275 (17.1) | 263 (16.3) | 0.020 |
| **Type 2 Diabetes Mellitus (%)** | | 2790 (27.6) | 461 (28.7) | 0.024 | 415 (25.8) | 461 (28.7) | 0.064 |
| **Cancer (%)** | | 2749 (27.2) | 480 (29.8) | 0.059 | 452 (28.1) | 480 (29.8) | 0.038 |
| **Hypertension (%)** | | 7808 (77.2) | 1193 (74.1) | 0.071 | 1174 (73.0) | 1193 (74.1) | 0.027 |
| **Substance Use Disorder (%)** | | 2938 (29.1) | 567 (35.2) | 0.133 | 558 (34.7) | 567 (35.2) | 0.012 |
| **Depression (%)** | | 3837 (37.9) | 682 (42.4) | 0.091 | 687 (42.7) | 682 (42.4) | 0.006 |
| **Hypertension Medication (%)** | | 6307 (62.4) | 1004 (62.4) | 0.001 | 1003 (62.3) | 1004 (62.4) | 0.001 |
| **T2DM Medication (%)** | | 1318 (13.0) | 225 (14.0) | 0.028 | 205 (12.7) | 225 (14.0) | 0.037 |
| **Lipid Lowering Medication (%)** | | 4646 (45.9) | 724 (45.0) | 0.019 | 713 (44.3) | 724 (45.0) | 0.014 |
| **Benzodiazepines Medication (%)** | | 36 (0.4) | 8 (0.5) | 0.022 | 8 (0.5) | 8 (0.5) | <0.001 |
| **Antidepressant Medication (%)** | | 5340 (52.8) | 871 (54.1) | 0.027 | 851 (52.9) | 871 (54.1) | 0.025 |
| **AD Medication (%)** | | 5339 (52.8) | 761 (47.3) | 0.110 | 760 (47.2) | 761 (47.3) | 0.001 |
| **Anticholinergics Medication (%)** | | 608 (6.0) | 75 (4.7) | 0.060 | 86 (5.3) | 75 (4.7) | 0.031 |
| **Analgesics Medication (%)** | | 878 (8.7) | 168 (10.4) | 0.060 | 152 (9.4) | 168 (10.4) | 0.033 |

Abbreviations: SMD, standardized mean difference; SD, standard deviation; T2DM, Type 2 Diabetes Mellitus; AD, Alzheimer’s Disease

**Supplementary Table 4 Baseline Characteristics of Patients Treated with Olanzapine and Risperidone Before and After Propensity Score Matching**

|  |  | **Before Propensity Score Matching** | | | **After Propensity Score Matching** | | |
| --- | --- | --- | --- | --- | --- | --- | --- |
|  |  | **Risperidone** | **Olanzapine** | **SMD** | **Risperidone** | **Olanzapine** | **SMD** |
| **n** |  | 2647 | 2313 |  | 2313 | 2313 |  |
| **Age (SD)** |  | 83.72 (8.30) | 82.98 (8.51) | 0.087 | 83.19 (8.35) | 82.98 (8.51) | 0.025 |
| **Sex (%)** | Female | 1690 (63.8) | 1422 (61.5) | 0.049 | 1450 (62.7) | 1422 (61.5) | 0.025 |
|  | Male | 957 (36.2) | 891 (38.5) |  | 863 (37.3) | 891 (38.5) |  |
| **Race (%)** | African American | 196 (7.4) | 143 (6.2) | 0.054 | 143 (6.2) | 143 (6.2) | <0.001 |
|  | Other | 112 (4.2) | 88 (3.8) |  | 88 (3.8) | 88 (3.8) |  |
|  | White | 2339 (88.4) | 2082 (90.0) |  | 2082 (90.0) | 2082 (90.0) |  |
| **Marital Status (%)** | Married | 886 (33.5) | 790 (34.2) | 0.028 | 786 (34.0) | 790 (34.2) | 0.010 |
|  | Other/Unknown | 928 (35.1) | 825 (35.7) |  | 818 (35.4) | 825 (35.7) |  |
|  | Unmarried | 833 (31.5) | 698 (30.2) |  | 709 (30.7) | 698 (30.2) |  |
| **Heart Failure (%)** | | 446 (16.8) | 420 (18.2) | 0.034 | 399 (17.3) | 420 (18.2) | 0.024 |
| **Myocardial Infarction (%)** | | 212 (8.0) | 216 (9.3) | 0.047 | 198 (8.6) | 216 (9.3) | 0.027 |
| **Stroke (%)** | | 402 (15.2) | 401 (17.3) | 0.058 | 376 (16.3) | 401 (17.3) | 0.029 |
| **Chronic Kidney Disease (%)** | | 703 (26.6) | 641 (27.7) | 0.026 | 629 (27.2) | 641 (27.7) | 0.012 |
| **Chronic Obstructive Pulmonary Disease (%)** | | 358 (13.5) | 340 (14.7) | 0.034 | 324 (14.0) | 340 (14.7) | 0.020 |
| **Type 2 Diabetes Mellitus (%)** | | 668 (25.2) | 658 (28.4) | 0.073 | 629 (27.2) | 658 (28.4) | 0.028 |
| **Cancer (%)** | | 671 (25.3) | 682 (29.5) | 0.093 | 643 (27.8) | 682 (29.5) | 0.037 |
| **Hypertension (%)** | | 1965 (74.2) | 1719 (74.3) | 0.002 | 1716 (74.2) | 1719 (74.3) | 0.003 |
| **Substance Use Disorder (%)** | | 758 (28.6) | 795 (34.4) | 0.124 | 735 (31.8) | 795 (34.4) | 0.055 |
| **Depression (%)** | | 1063 (40.2) | 982 (42.5) | 0.047 | 966 (41.8) | 982 (42.5) | 0.014 |
| **Hypertension Medication (%)** | | 1569 (59.3) | 1437 (62.1) | 0.058 | 1428 (61.7) | 1437 (62.1) | 0.008 |
| **T2DM Medication (%)** | | 338 (12.8) | 327 (14.1) | 0.040 | 309 (13.4) | 327 (14.1) | 0.023 |
| **Lipid Lowering Medication (%)** | | 1142 (43.1) | 1063 (46.0) | 0.057 | 1049 (45.4) | 1063 (46.0) | 0.012 |
| **Benzodiazepines Medication (%)** | | 8 (0.3) | 13 (0.6) | 0.040 | 7 (0.3) | 13 (0.6) | 0.040 |
| **Antidepressant Medication (%)** | | 1479 (55.9) | 1279 (55.3) | 0.012 | 1277 (55.2) | 1279 (55.3) | 0.002 |
| **AD Medication (%)** | | 1431 (54.1) | 1136 (49.1) | 0.099 | 1178 (50.9) | 1136 (49.1) | 0.036 |
| **Anticholinergics Medication (%)** | | 131 (4.9) | 118 (5.1) | 0.007 | 116 (5.0) | 118 (5.1) | 0.004 |
| **Analgesics Medication (%)** | | 211 (8.0) | 233 (10.1) | 0.073 | 201 (8.7) | 233 (10.1) | 0.047 |

Abbreviations: SMD, standardized mean difference; SD, standard deviation; T2DM, Type 2 Diabetes Mellitus; AD, Alzheimer’s Disease

**Supplementary Table 5 Baseline Characteristics of Patients Treated with Risperidone and Quetiapine Before and After Propensity Score Matching**

|  |  | **Before Propensity Score Matching** | | | **After Propensity Score Matching** | | |
| --- | --- | --- | --- | --- | --- | --- | --- |
|  |  | **Quetiapine** | **Risperidone** | **SMD** | **Quetiapine** | **Risperidone** | **SMD** |
| **n** |  | 10097 | 1914 |  | 1914 | 1914 |  |
| **Age (SD)** |  | 84.33 (8.01) | 83.79 (8.49) | 0.066 | 83.94 (8.31) | 83.79 (8.49) | 0.018 |
| **Sex (%)** | Female | 6192 (61.3) | 1245 (65.0) | 0.077 | 1290 (67.4) | 1245 (65.0) | 0.050 |
|  | Male | 3905 (38.7) | 669 (35.0) |  | 624 (32.6) | 669 (35.0) |  |
| **Race (%)** | African American | 686 (6.8) | 142 (7.4) | 0.028 | 143 (7.5) | 142 (7.4) | 0.013 |
|  | Other | 446 (4.4) | 79 (4.1) |  | 84 (4.4) | 79 (4.1) |  |
|  | White | 8965 (88.8) | 1693 (88.5) |  | 1687 (88.1) | 1693 (88.5) |  |
| **Marital Status (%)** | Married | 3591 (35.6) | 600 (31.3) | 0.099 | 578 (30.2) | 600 (31.3) | 0.034 |
|  | Other/Unknown | 3452 (34.2) | 663 (34.6) |  | 655 (34.2) | 663 (34.6) |  |
|  | Unmarried | 3054 (30.2) | 651 (34.0) |  | 681 (35.6) | 651 (34.0) |  |
| **Heart Failure (%)** | | 1829 (18.1) | 324 (16.9) | 0.031 | 333 (17.4) | 324 (16.9) | 0.012 |
| **Myocardial Infarction (%)** | | 969 (9.6) | 159 (8.3) | 0.045 | 163 (8.5) | 159 (8.3) | 0.008 |
| **Stroke (%)** | | 1660 (16.4) | 274 (14.3) | 0.059 | 285 (14.9) | 274 (14.3) | 0.016 |
| **Chronic Kidney Disease (%)** | | 2980 (29.5) | 524 (27.4) | 0.047 | 501 (26.2) | 524 (27.4) | 0.027 |
| **Chronic Obstructive Pulmonary Disease (%)** | | 1373 (13.6) | 272 (14.2) | 0.018 | 259 (13.5) | 272 (14.2) | 0.020 |
| **Type 2 Diabetes Mellitus (%)** | | 2808 (27.8) | 486 (25.4) | 0.055 | 457 (23.9) | 486 (25.4) | 0.035 |
| **Cancer (%)** | | 2760 (27.3) | 470 (24.6) | 0.063 | 481 (25.1) | 470 (24.6) | 0.013 |
| **Hypertension (%)** | | 7795 (77.2) | 1416 (74.0) | 0.075 | 1404 (73.4) | 1416 (74.0) | 0.014 |
| **Substance Use Disorder (%)** | | 2943 (29.1) | 534 (27.9) | 0.028 | 510 (26.6) | 534 (27.9) | 0.028 |
| **Depression (%)** | | 3873 (38.4) | 792 (41.4) | 0.062 | 774 (40.4) | 792 (41.4) | 0.019 |
| **Hypertension Medication (%)** | | 6325 (62.6) | 1139 (59.5) | 0.064 | 1136 (59.4) | 1139 (59.5) | 0.003 |
| **T2DM Medication (%)** | | 1328 (13.2) | 242 (12.6) | 0.015 | 227 (11.9) | 242 (12.6) | 0.024 |
| **Lipid Lowering Medication (%)** | | 4634 (45.9) | 789 (41.2) | 0.094 | 782 (40.9) | 789 (41.2) | 0.007 |
| **Benzodiazepines Medication (%)** | | 39 (0.4) | 6 (0.3) | 0.012 | 9 (0.5) | 6 (0.3) | 0.025 |
| **Antidepressant Medication (%)** | | 5374 (53.2) | 1095 (57.2) | 0.080 | 1082 (56.5) | 1095 (57.2) | 0.014 |
| **AD Medication (%)** | | 5315 (52.6) | 1025 (53.6) | 0.018 | 1029 (53.8) | 1025 (53.6) | 0.004 |
| **Anticholinergics Medication (%)** | | 609 (6.0) | 93 (4.9) | 0.052 | 105 (5.5) | 93 (4.9) | 0.028 |
| **Analgesics Medication (%)** | | 903 (8.9) | 165 (8.6) | 0.011 | 172 (9.0) | 165 (8.6) | 0.013 |

Abbreviations: SMD, standardized mean difference; SD, standard deviation; T2DM, Type 2 Diabetes Mellitus; AD, Alzheimer’s Disease

**Supplementary Table 6 Summary Table of All the Sensitivity Analyses**

| **Comparison pairs**  **Cox model parameter setting** | **2yr follow-up, 30 days grace period**  **(result in the main text)** | **2yr follow-up, 14 days grace period** | **2yr follow-up, 60 days grace period** | **2yr follow-up, 90 days grace period** | **1yr follow-up, 30 days grace period** |
| --- | --- | --- | --- | --- | --- |
| **Aripiprazole vs. Olanzapine** | HR = 0.667,  p = 0.0212 | HR = 0.583,  p = 0.00726 | HR = 0.741,  p = 0.0442 | HR = 0.703,  p = 0.0117 | HR = 0.683,  p = 0.0306 |
| **Aripiprazole vs. Quetiapine** | HR = 0.677,  p = 0.044 | HR = 0.8,  p = 0.296 | HR = 0.688,  p = 0.0212 | HR = 0.691,   p = 0.0148 | HR = 0.697,   p = 0.064 |
| **Aripiprazole vs. Risperidone** | HR = 0.75,  p = 0.114 | HR = 0.815,  p = 0.316 | HR = 0.789,  p = 0.11 | HR = 0.677,   p = 0.00447 | HR = 0.75,  p = 0.114 |
| **Quetiapine vs. Olanzapine** | HR = 0.833,  p = 0.0378 | HR = 0.824,  p = 0.0451 | HR = 0.795,  p = 0.00298 | HR = 0.773,  p = 0.000445 | HR = 0.818,  p = 0.0235 |
| **Risperidone vs. Olanzapine** | HR = 0.869,  p = 0.073 | HR = 0.873,  p = 0.13 | HR = 0.887,  p = 0.079 | HR = 0.940,  p = 0.32 | HR = 0.872,  p = 0.084 |
| **Quetiapine vs. Risperidone** | HR = 0.830,  p = 0.0255 | HR = 0.894,  p = 0.227 | HR = 0.886,  p = 0.102 | HR = 0.876,  p = 0.0546 | HR = 0.841,  p = 0.0378 |

**Supplementary Table 7** Subgroup Cox Analyses Results Based on HTE-defined Subgroups

| **Comparison Pair** | **HR** | **95% CI Lower** | **95% CI Upper** | **p-value** |
| --- | --- | --- | --- | --- |
| Aripiprazole vs. Quetiapine (T2DM users) | 1 | 0.3225 | 3.101 | NA |
| Aripiprazole vs. Risperidone (T2DM users) | 0.375 | 0.1338 | 1.051 | 0.0621 |
| Aripiprazole vs. Quetiapine + Risperidone (T2DM users) | 0.6038 | 0.4386 | 0.8311 | 0.00197 |
| Quetiapine vs. Olanzapine (AD medication users) | 0.8196 | 0.6285 | 1.069 | 0.142 |
| Aripiprazole vs. Olanzapine (AD medication users) | 0.6111 | 0.3541 | 1.055 | 0.0769 |
| Quetiapine + Aripiprazole vs. Olanzapine (AD medication users) | 0.911 | 0.737 | 1.1259 | 0.389 |

**Supplementary Figure 1** Kaplan-Meier curves for all-cause mortality among patients treated with aripiprazole versus olanzapine.
(A) Unadjusted survival curves based on raw data.
(B) Survival curves after 1:1 propensity score matching.
(C) Survival curves in the matched cohort, limited to 2-year follow-up.

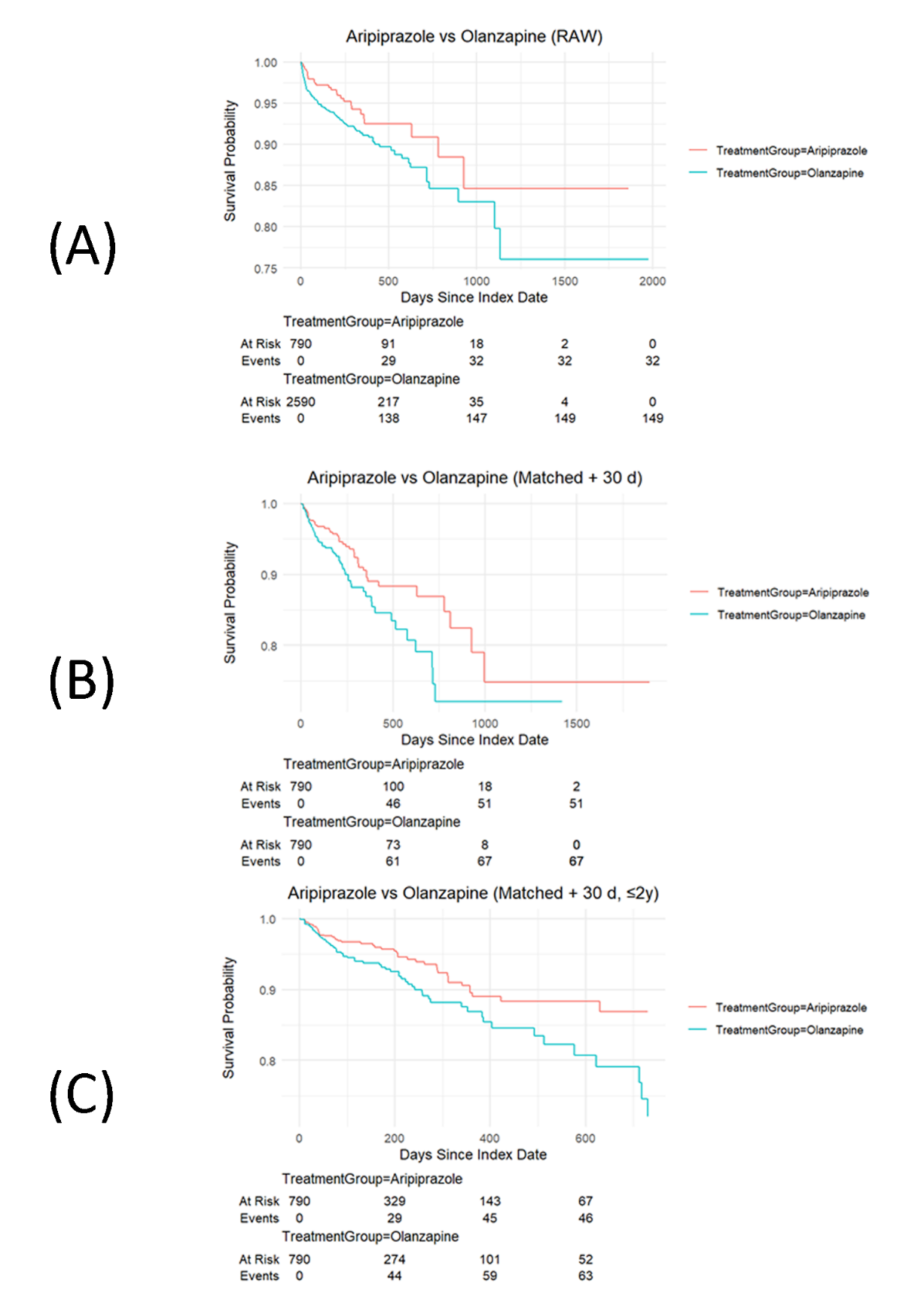

**Supplementary Figure 2** Kaplan-Meier curves for all-cause mortality among patients treated with aripiprazole versus quetiapine.
(A) Unadjusted survival curves based on raw data.
(B) Survival curves after 1:1 propensity score matching.
(C) Survival curves in the matched cohort, limited to 2-year follow-up.

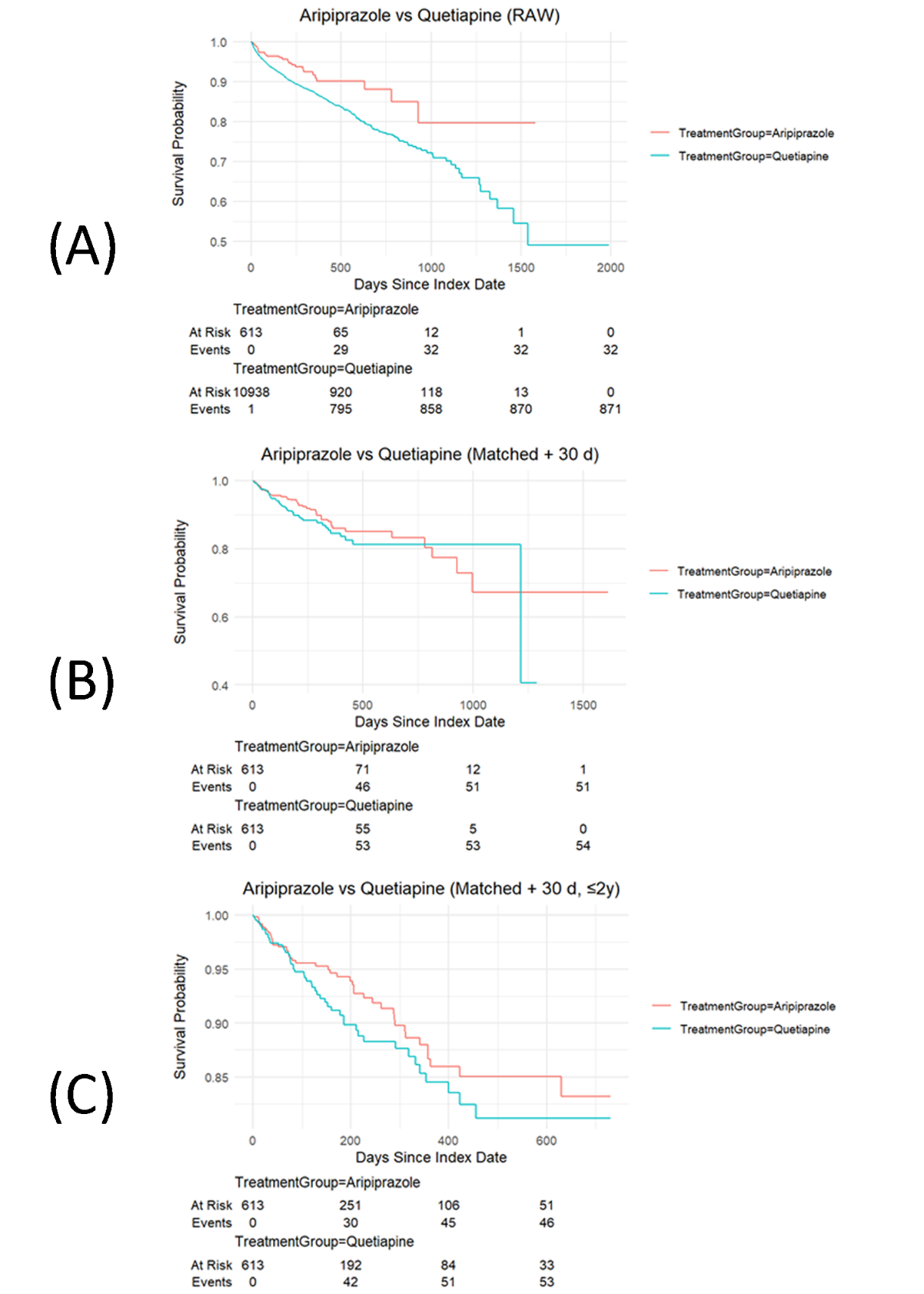

**Supplementary Figure 3** Kaplan-Meier curves for all-cause mortality among patients treated with aripiprazole versus risperidone.
(A) Unadjusted survival curves based on raw data.
(B) Survival curves after 1:1 propensity score matching.
(C) Survival curves in the matched cohort, limited to 2-year follow-up.

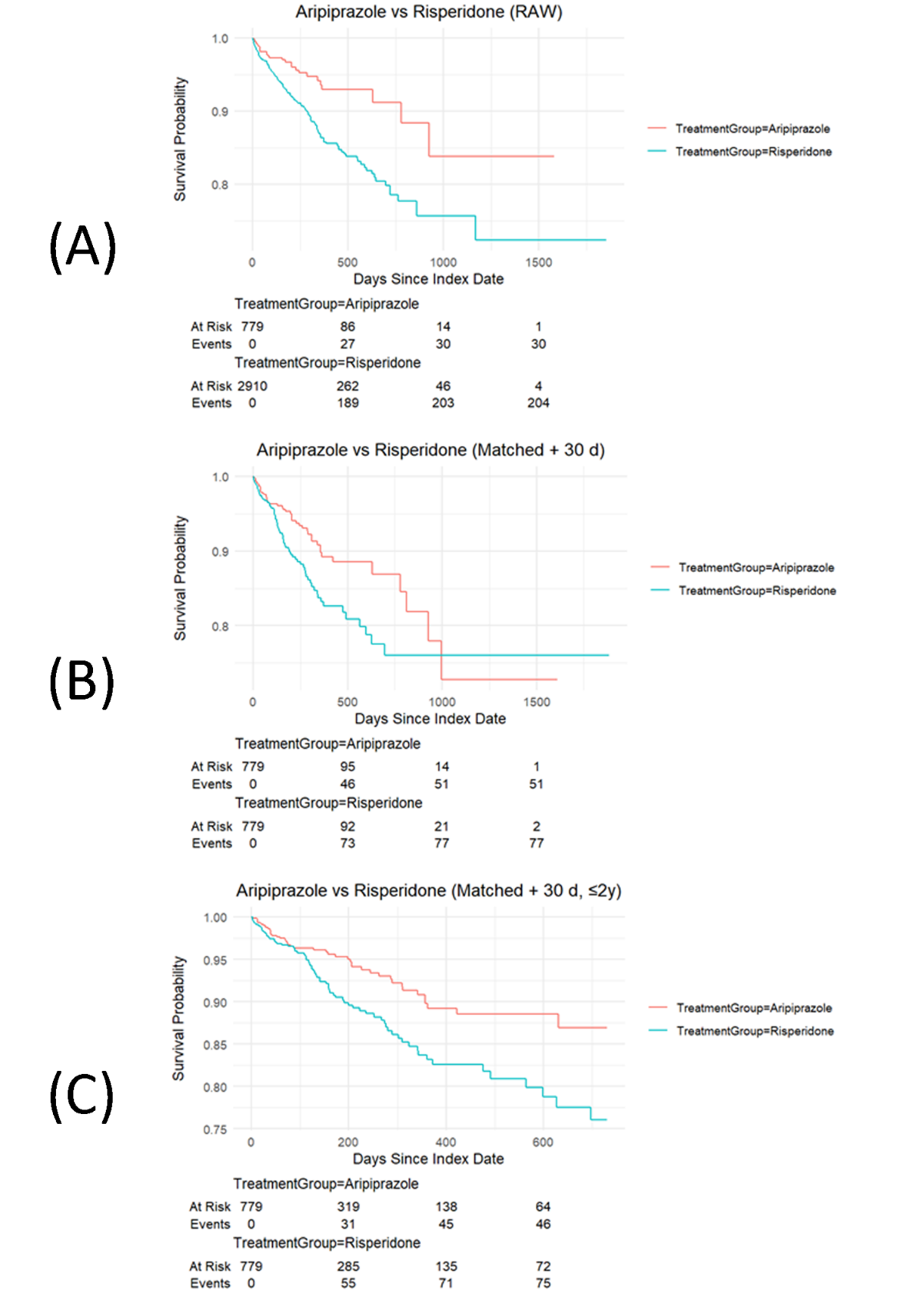

**Supplementary Figure 4** Kaplan-Meier curves for all-cause mortality among patients treated with olanzapine versus quetiapine.
(A) Unadjusted survival curves based on raw data.
(B) Survival curves after 1:1 propensity score matching.
(C) Survival curves in the matched cohort, limited to 2-year follow-up.

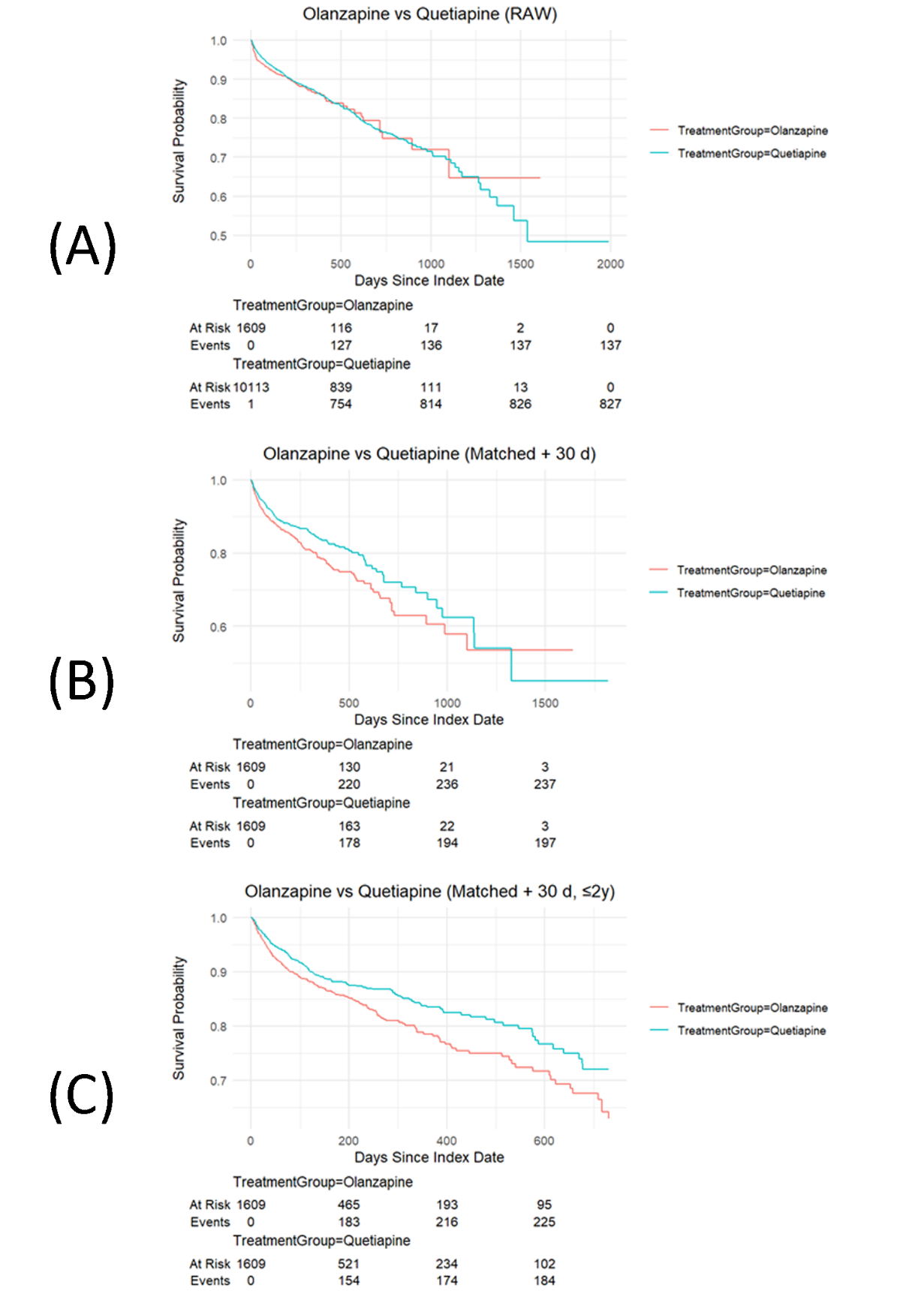

**Supplementary Figure 5** Kaplan-Meier curves for all-cause mortality among patients treated with olanzapine versus risperidone.
(A) Unadjusted survival curves based on raw data.
(B) Survival curves after 1:1 propensity score matching.
(C) Survival curves in the matched cohort, limited to 2-year follow-up.

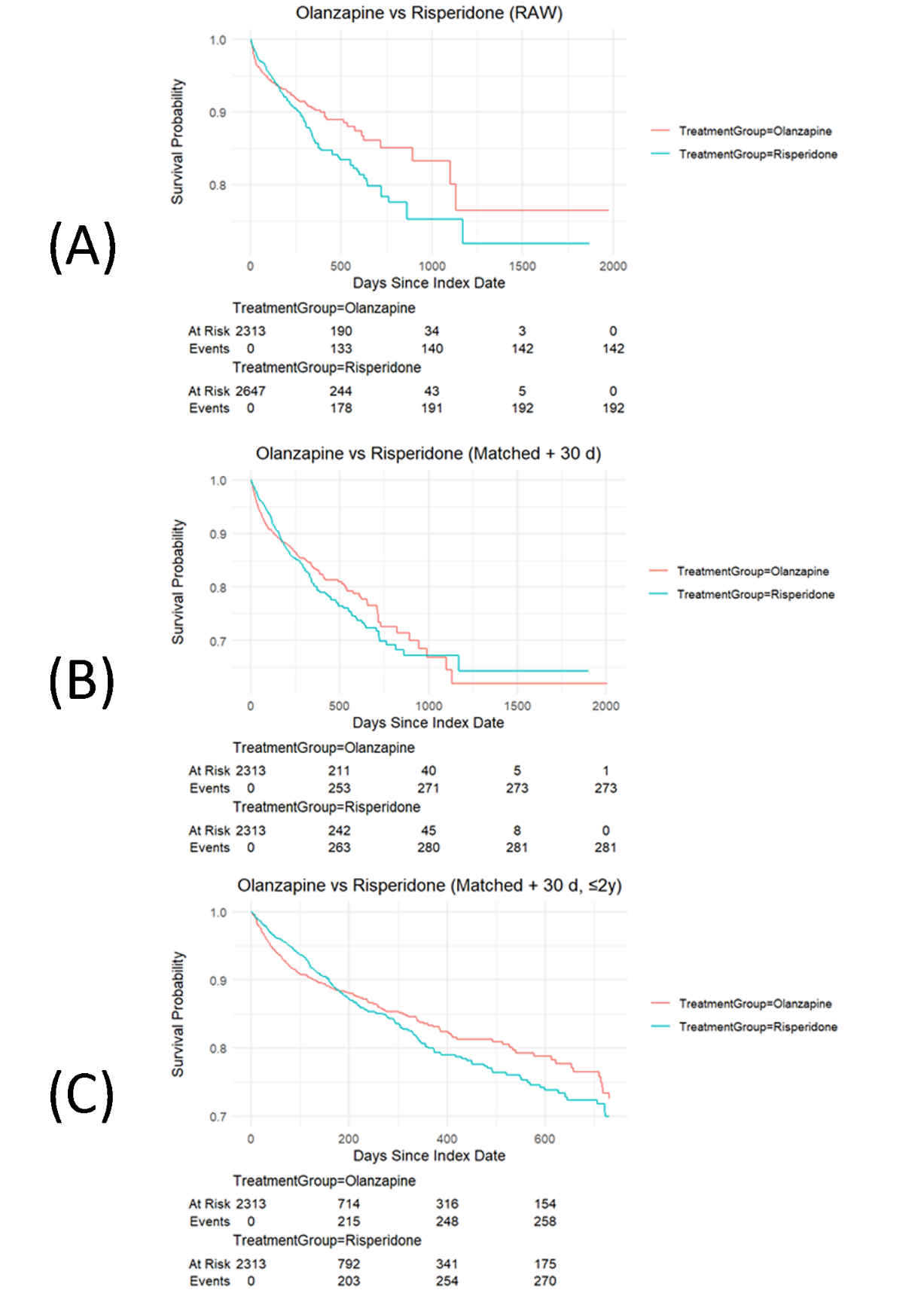

**Supplementary Figure 6** Kaplan-Meier curves for all-cause mortality among patients treated with risperidone versus quetiapine.
(A) Unadjusted survival curves based on raw data.
(B) Survival curves after 1:1 propensity score matching.
(C) Survival curves in the matched cohort, limited to 2-year follow-up.

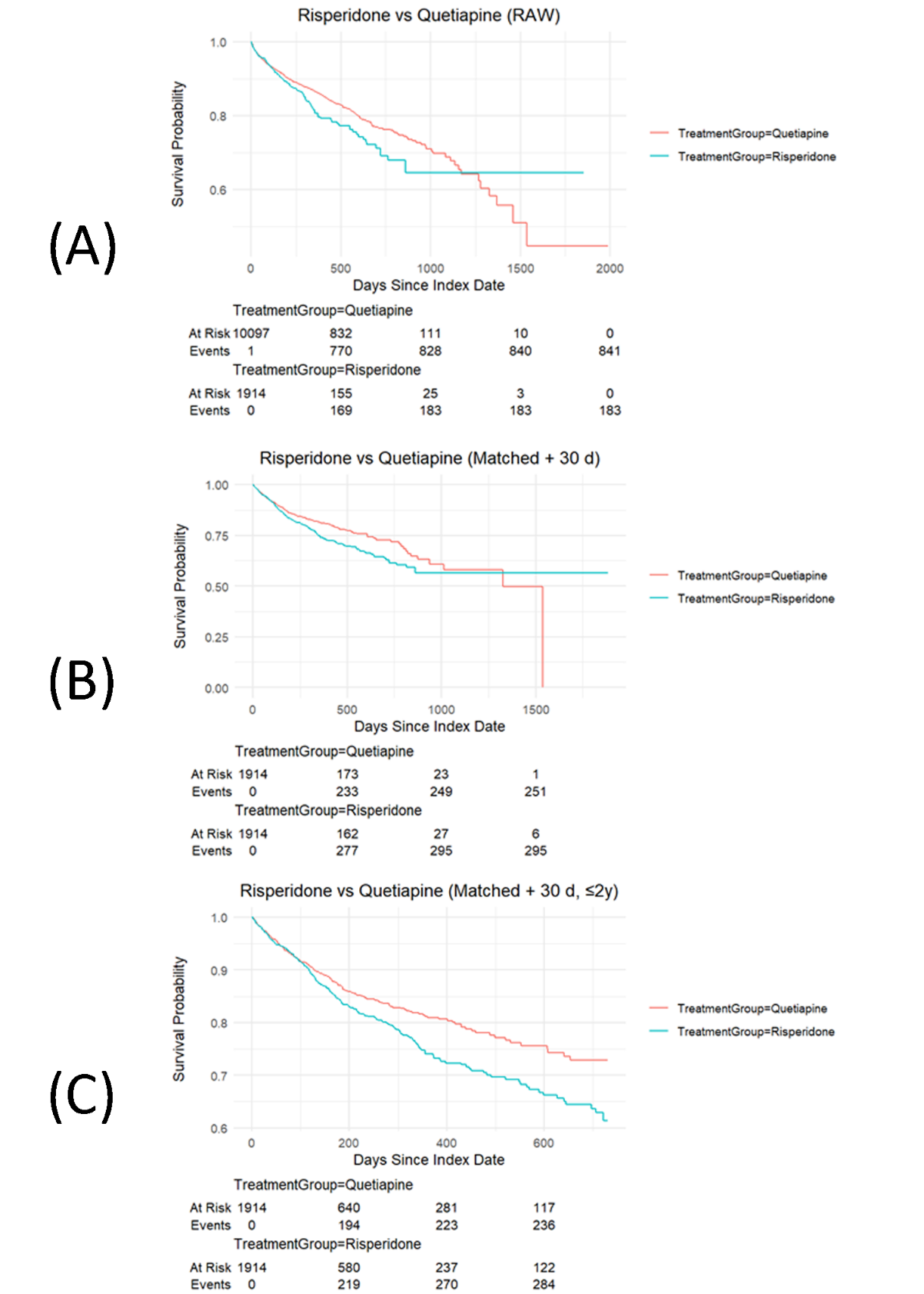

**Supplementary Figure 7** Subgroups with heterogeneous treatment effects on all-cause mortality among aripiprazole vs. olanzapine.

Causal tree analysis identified patient subgroups with significantly different average treatment effects (ATEs) of aripiprazole compared to olanzapine. Terminal nodes represent subgroups, with corresponding ATE values. Negative ATEs indicate reduced mortality risk associated with aripiprazole. All estimates were derived from the matched analytic sample. Binary covariates (e.g., T2DM Medication; Depression) are coded as 1 = presence/yes and 0 = absence/no.

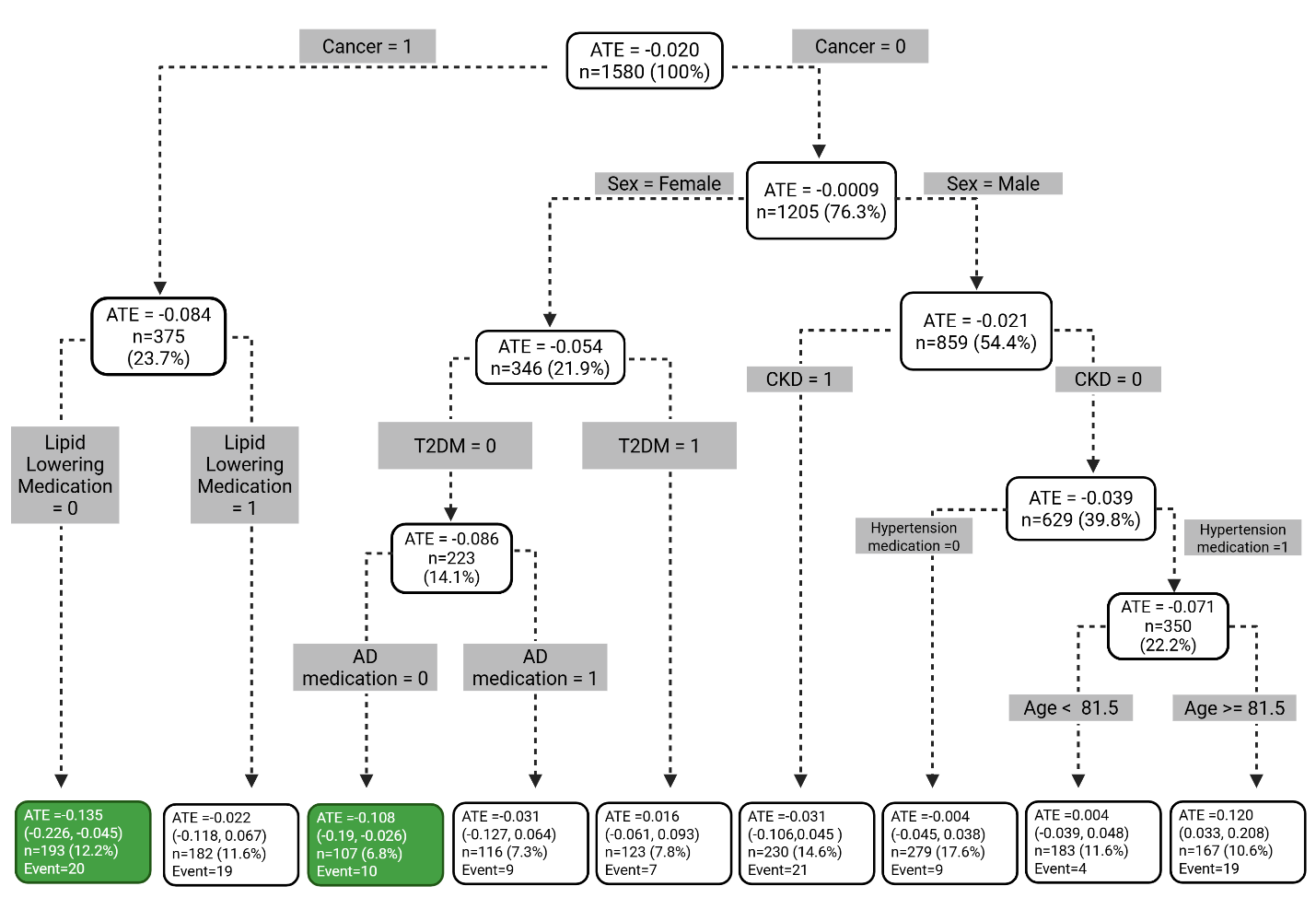

**Supplementary Figure 8** Subgroups with heterogeneous treatment effects on all-cause mortality among olanzapine vs. quetiapine.

Causal tree analysis identified patient subgroups with significantly different average treatment effects (ATEs) of olanzapine compared to quetiapine. Terminal nodes represent subgroups, with corresponding ATE values. Negative ATEs indicate reduced mortality risk associated with olanzapine. All estimates were derived from the matched analytic sample. Binary covariates (e.g., T2DM Medication; Depression) are coded as 1 = presence/yes and 0 = absence/no.

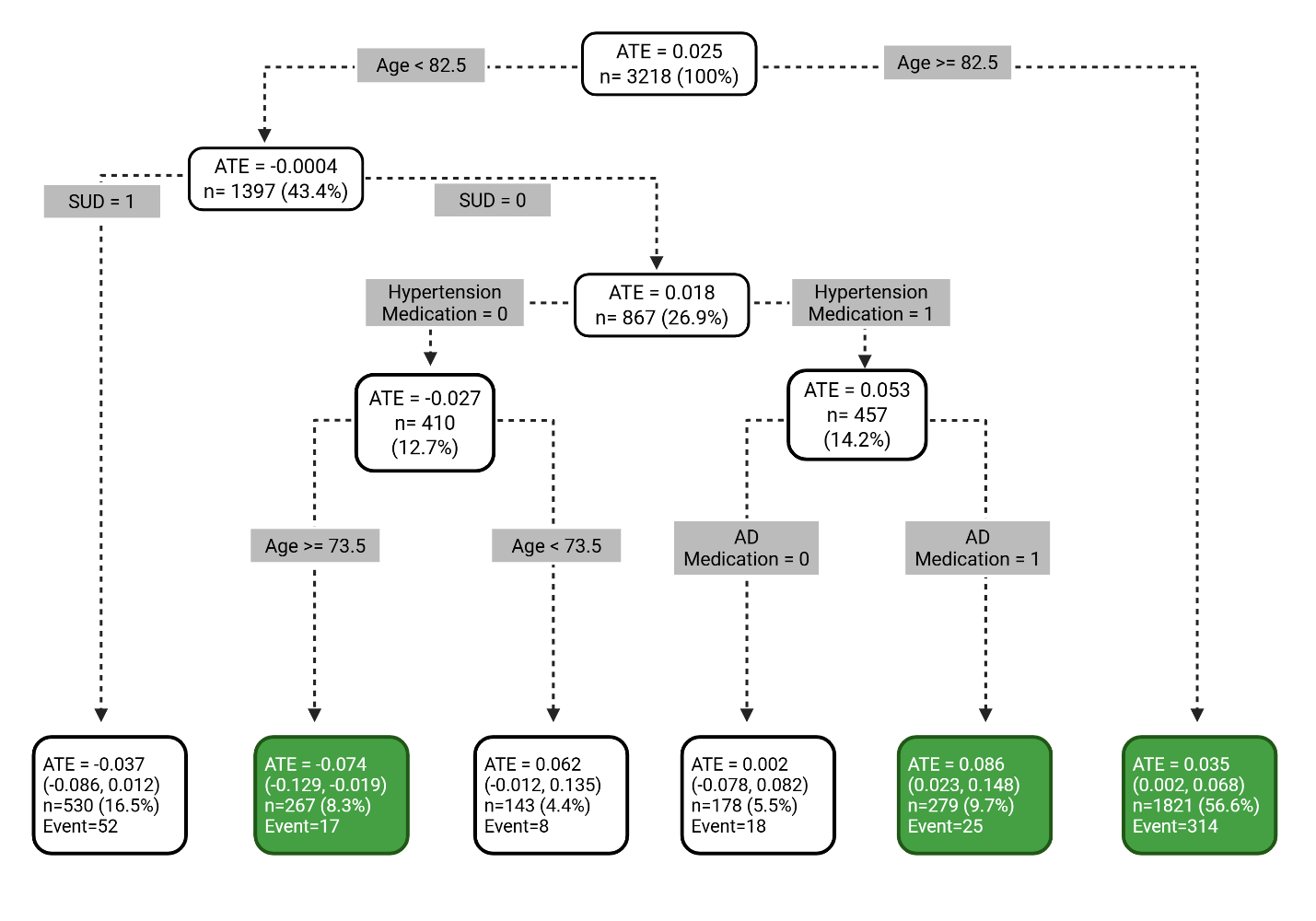

**Supplementary Figure 9 Subgroups with heterogeneous treatment effects on all-cause mortality among olanzapine vs. risperidone.**
Causal tree analysis identified patient subgroups with significantly different average treatment effects (ATEs) of olanzapine compared to risperidone. Terminal nodes represent subgroups, with corresponding ATE values. Negative ATEs indicate reduced mortality risk associated with olanzapine. All estimates were derived from the matched analytic sample. Binary covariates (e.g., T2DM Medication; Depression) are coded as 1 = presence/yes and 0 = absence/no.

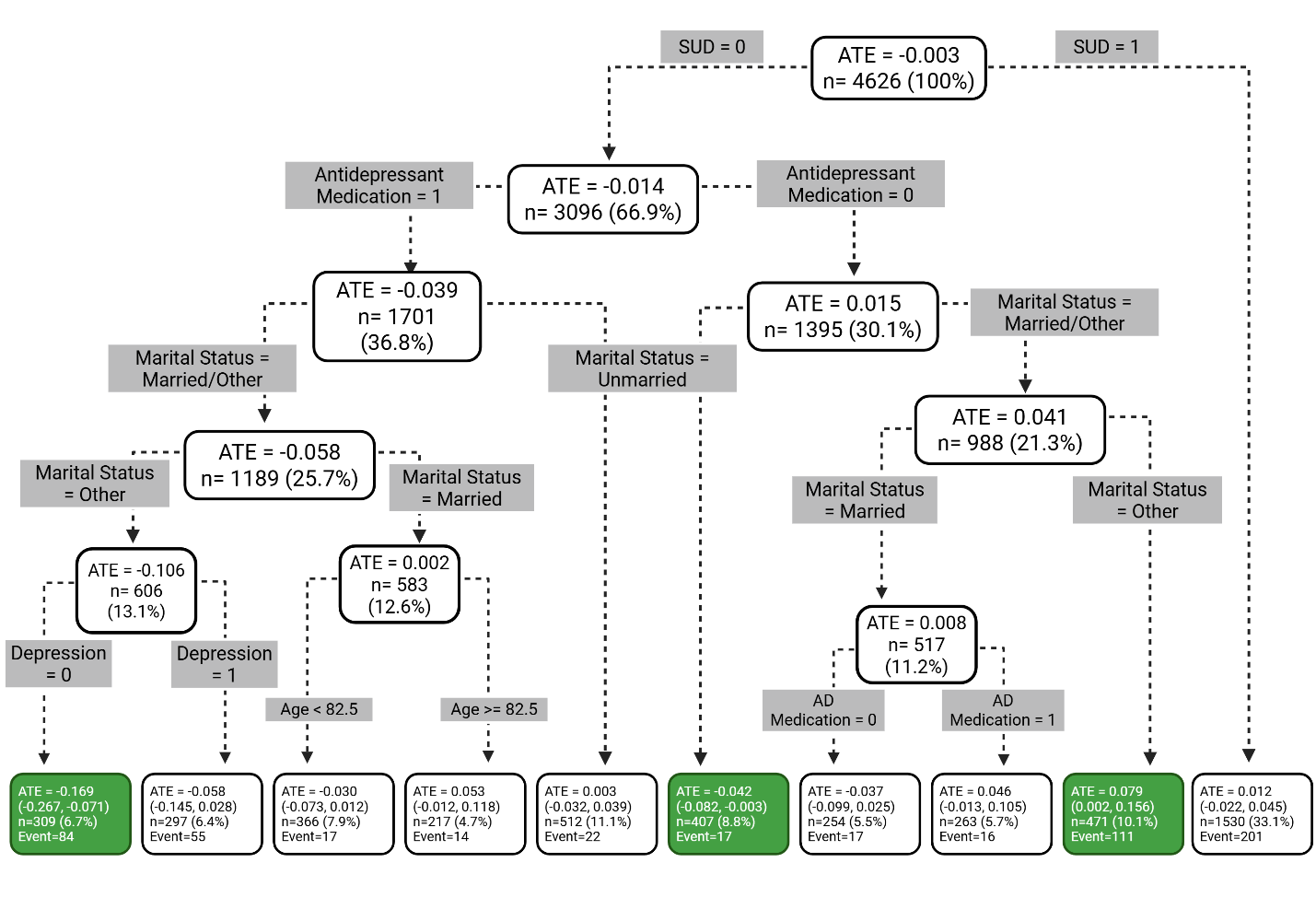

**Supplementary Figure 10** Subgroups with heterogeneous treatment effects on all-cause mortality among risperidone vs. quetiapine.

Causal tree analysis identified patient subgroups with significantly different average treatment effects (ATEs) of risperidone compared to quetiapine. Terminal nodes represent subgroups, with corresponding ATE values. Negative ATEs indicate reduced mortality risk associated with risperidone. All estimates were derived from the matched analytic sample. Binary covariates (e.g., T2DM Medication; Depression) are coded as 1 = presence/yes and 0 = absence/no.

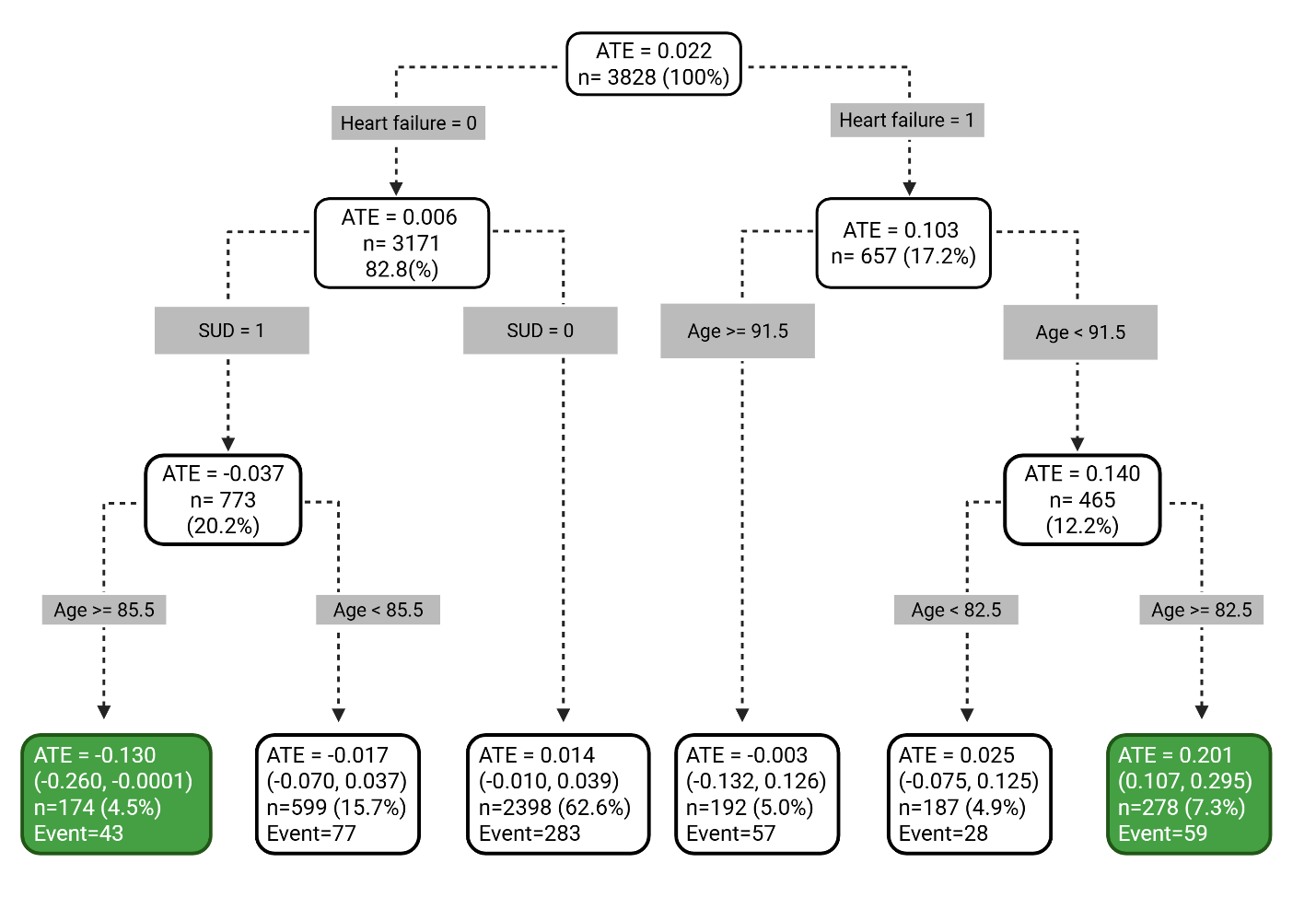
